# Network Reorganization of Anterograde Thalamic Connectivity During Non-Periodic Patterned Deep Brain Stimulation

**DOI:** 10.64898/2026.01.22.26344468

**Authors:** Teryn D. Johnson, Bobby Mohan, Harvey Huang, Justin Cramer, Cornelia Drees, Matthew Hoerth, Amy Crepeau, Joseph Drazkowski, Katherine Noe, Leslie Baxter, Amir A. Mbonde, Christopher Harris, Nuri Ince, Kai Miller, Dora Hermes, Gregory Worrell, Jonathon J. Parker

## Abstract

Deep brain stimulation (DBS) of the anterior nucleus of the thalamus (ANT) can reduce seizures in patients with drug-resistant epilepsy. However, seizure freedom is rare, and there is no early response biomarker to predict long-term seizure suppression. To evaluate a short-term biomarker of therapeutic response to DBS, we studied 18-minute trials of ANT stimulation in patients with drug-resistant epilepsy undergoing intracranial electroencephalography. We compared a standard high-frequency periodic versus a non-periodic pulse pattern. We found that short-term ANT stimulation within the clinically used amplitude range preferentially altered the first 150 ms of anterograde ANT to anterior cingulate and superior frontal connectivity. Non-periodic ANT stimulation selectively modified early cortico-cortical evoked potentials more than periodic stimulation, and these changes were driven by the response phase. These pattern-specific changes in connectivity between the ANT and spatially restricted cortical areas suggest differing mechanisms and are a promising biomarker for patients being treated with ANT-DBS.

---

Deep brain stimulation (DBS) of the anterior nucleus of the thalamus (ANT) is a Food and Drug Administration-approved therapy for reducing seizure frequency in patients with focal-onset, limbic network, drug-resistant epilepsy^1^. For patients that either cannot be treated with surgical resection, or their resection has failed to stop seizures, DBS remains one of only a few available options. Although DBS can reduce seizures^2,3^, its outcomes vary widely and rarely result in complete seizure freedom^1–6^. To explain these variable outcomes, researchers have evaluated whether placement variance of the ANT-DBS lead contributes to suboptimal treatment. They found final lead location accounts for only part of the variability in seizure rate^7,8^, suggesting that clinical outcomes may also be related to therapy-specific or patient-intrinsic factors including stimulation parameters or seizure network specific constraints.

To explore variability in patient outcomes to ANT-DBS, common approaches focus on changes to neural activity in either the ANT or the patient’s seizure-onset zone (SOZ). However, this approach may be too limited in scope, with many studies supporting that epilepsy is a network disorder involving more than just a single aberrant brain structure. For example, in intracranial electroencephalogram (EEG) and resting-state functional magnetic resonance imaging (fMRI) studies, patients with epilepsy have widespread connectivity changes^9,10^. During ANT-DBS, fMRI-based characterization of macroscale blood-flow patterns has suggested that patient-specific connectivity—particularly within non-epileptic networks—may influence seizure control^11–14^. Further, long-term brain network reorganization has been associated with seizure suppression in patients undergoing responsive neuromodulation^15^. In humans and non-human primates, direct intracranial and transcranial brain stimulation alters cortical connectivity and excitability^16–18^. Lastly, emerging evidence supports that ANT-DBS can change the response in connected structures after longer timescales of stimulation (hours) and is associated with interictal discharge suppresion^19^. These converging findings support that (1) epilepsy can be characterized by abnormal network connectivity, (2) brain stimulation modifies region-to-region communication within the brain, and (3) these modifications may be related to long-term seizure outcome.

To improve therapeutic DBS outcomes, researchers must address two important issues in epilepsy research: clinical heterogeneity and the intrinsic delay in evaluating treatment success. Even among patients with similar SOZs, seizure propagation and thalamic recruitment can vary significantly^20,21^ leading to significant clinical heterogeneity. With this heterogeneity in brain networks between patients, understanding how DBS affects distinct networks is crucial. Decades of clinical treatment has made it clear that DBS must be optimized per patient, which is a slow and iterative process that often requires months to years of parameter tuning to significantly reduce seizures^22,23^. In terms of intrinsic delays, unlike many symptoms of movement disorders that show immediate effects after DBS^24^, seizure reduction unfolds over months. Adding to the further difficulty of DBS parameter optimization, these seizure rates are measured by patient reports, which are imprecise^25,26^ and confounded by seizure cycles that fluctuate over days and weeks^27–29^. To improve DBS therapy for the treatment of epilepsy, clinicians need an objective biomarker that (1) is detectable in smaller time windows, (2) predicts long-term efficacy, and (3) can be measured during a routine programming session.

Beyond the intrinsic challenges of current methods of DBS parameter optimization, it is also not clear which parameters most critically need adjustments to optimally reduce seizure rates. Typically, clinical protocols for brain-directed neuromodulation focus on adjusting current and frequency^22^, but altering the stimulation pattern itself may improve seizure suppression^30–35^. Patterned brain stimulation, such as theta-burst brain stimulation, has demonstrated region-specific effects on brain networks^36^. However, it is currently underexplored how stimulation patterns impact connectivity between networks. Traditional periodic stimulation delivers evenly spaced pulses that produce a narrow frequency profile, whereas non-periodic stimulation introduces timing jitter that produces a broader frequency range. This temporal complexity may engage more diverse neural populations and reduce pathological synchrony^33,34,37^.

In this study, we aimed to evaluate a short-term biomarker of therapeutic response to DBS by examining how ANT stimulation modifies connectivity using cortico-cortical evoked potentials (CCEPs). Although we applied stimulation subcortically in the ANT, we use the term *CCEP* because it is commonly recognized in the field to refer to cortically recorded potentials evoked by single-pulse electrical stimulation. CCEPs correlate well with diffusion tensor imaging measures of structural connectivity^38^.

Because CCEPs uniquely capture effective connectivity^39–43^, they are ideally suited to detect connectivity changes arising from therapeutic stimulation^43,44^. We measured CCEPs before and after ANT stimulation in patients with drug-resistant epilepsy undergoing intracranial monitoring. While exploring how ANT-DBS affects network connectivity, we evaluated whether parameter-related differences in current (high vs low amplitude) and pattern (non-periodic vs periodic stimulation) were detectable and contributed to network changes.

## RESULTS

### ANT stimulation preferentially modifies anterograde thalamo-frontal connectivity

The ANT is a hub in the limbic network with broad connectivity to both subcortical and cortical regions^45,46^. We hypothesized that short-term ANT stimulation could induce broad changes in CCEPs within the limbic network. After intracranial monitoring for seizure localization, patients (n = 9) underwent 18-minute stimulation blocks in the ANT, interspersed with single-pulse stimuli to measure CCEPs (Fig. 1a,b). We recorded signals using stereoelectroencephalography (SEEG) electrodes and compared modifications before vs after DBS-like therapeutic stimulation. These modifications are referred to as response modifications throughout this paper when referring to specific differences between the evoked waveforms. When we generalize the response results across recording channels and across the cohort, we instead refer to these modifications as connectivity modifications as a more general term highlighting the theoretical underpinning of the phenomenon behind the modified response.

**Fig. 1:**
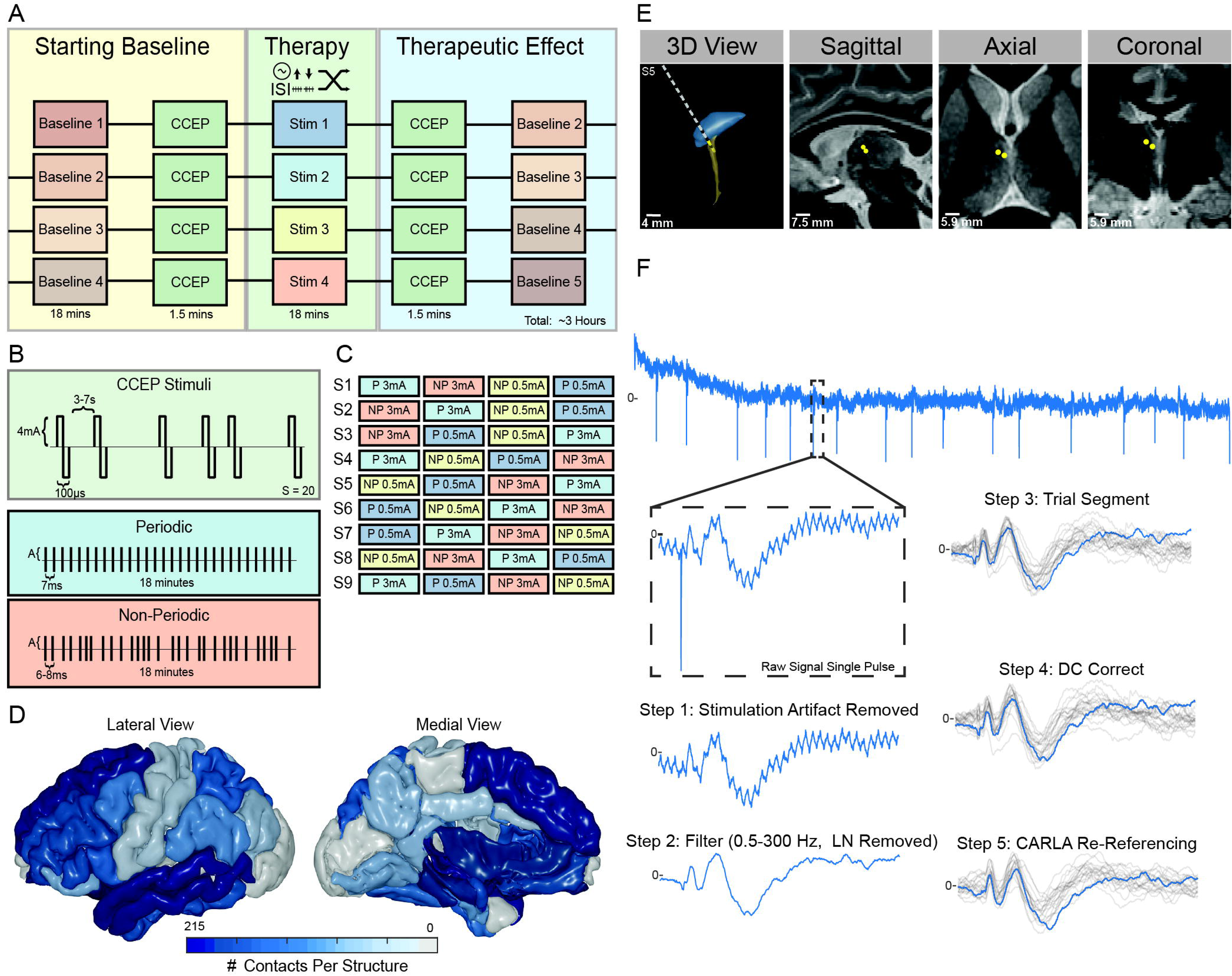
Thalamic-trial stimulation paradigm and iEEG data preprocessing workflow. **a**, iEEG data and CCEPs were obtained before and after short periods (18 minutes) of trial thalamic stimulation via intracranial electrodes. **b**, Illustration of stimulation protocols: single-pulse CCEP stimuli, periodic stimulation, and non-periodic (randomly jittered interval) stimulation. **c**, Stimulation sequence for periodic (P) and non-periodic (NP) patterns and current amplitude (0.5 or 3 mA) were pseudo-randomized across patients in the cohort (n = 9) to minimize ordering effects. **d**, Heat map of electrode contact locations across all patients, collapsed across hemispheres. Color hue indicates the total sum of recording contacts across the patient cohort per anatomic structure according to FreeSurfer brain parcellation using DKTatlas. **e**, The ANT-MTT junction was targeted for clinical purposes during intracranial monitoring. Images include a three-dimensional (3D) visualization of electrode placement and representative sagittal, axial, and coronal fGATIR MRI reconstructions. **f**, Overview of preprocessing steps applied to raw iEEG data acquired during single-pulse stimulation: (1) removal of stimulation artifact, (2) filtering (0.5–300 Hz, line noise [LN] removed), (3) trial segmentation around stimuli, (4) DC correction via baseline subtraction, and (5) Common Average Referencing using a Localized Adaptive Reference scheme (CARLA).

We manipulated two key stimulation parameters: current amplitude (0.5 mA “low” vs 3 mA “high”) and stimulation pattern (non-periodic vs periodic). We delivered biphasic pulses through the two electrode contacts closest to the ANT-mammillothalamic tract (MTT) junction (Fig. 1e, Supplementary Fig. 1a,b). High-current stimulation markedly changed CCEPs, whereas low-current stimulation produced negligible differences (see representative tracing in Fig. 2b,c). Across all patients, the percentage of post-pulse time points (15–500 ms) with significant changes was 1.05% after high-current stimulation, but only 0.01% after low-current stimulation following Bonferroni significance threshold correction for multiple comparisons (Fig. 3d). High-current stimulation was more associated with connectivity modifications (χ^2^ = 33228.56, *P* < 0.00001). Consequently, we focused most analyses on the high-current condition.

**Fig. 2:**
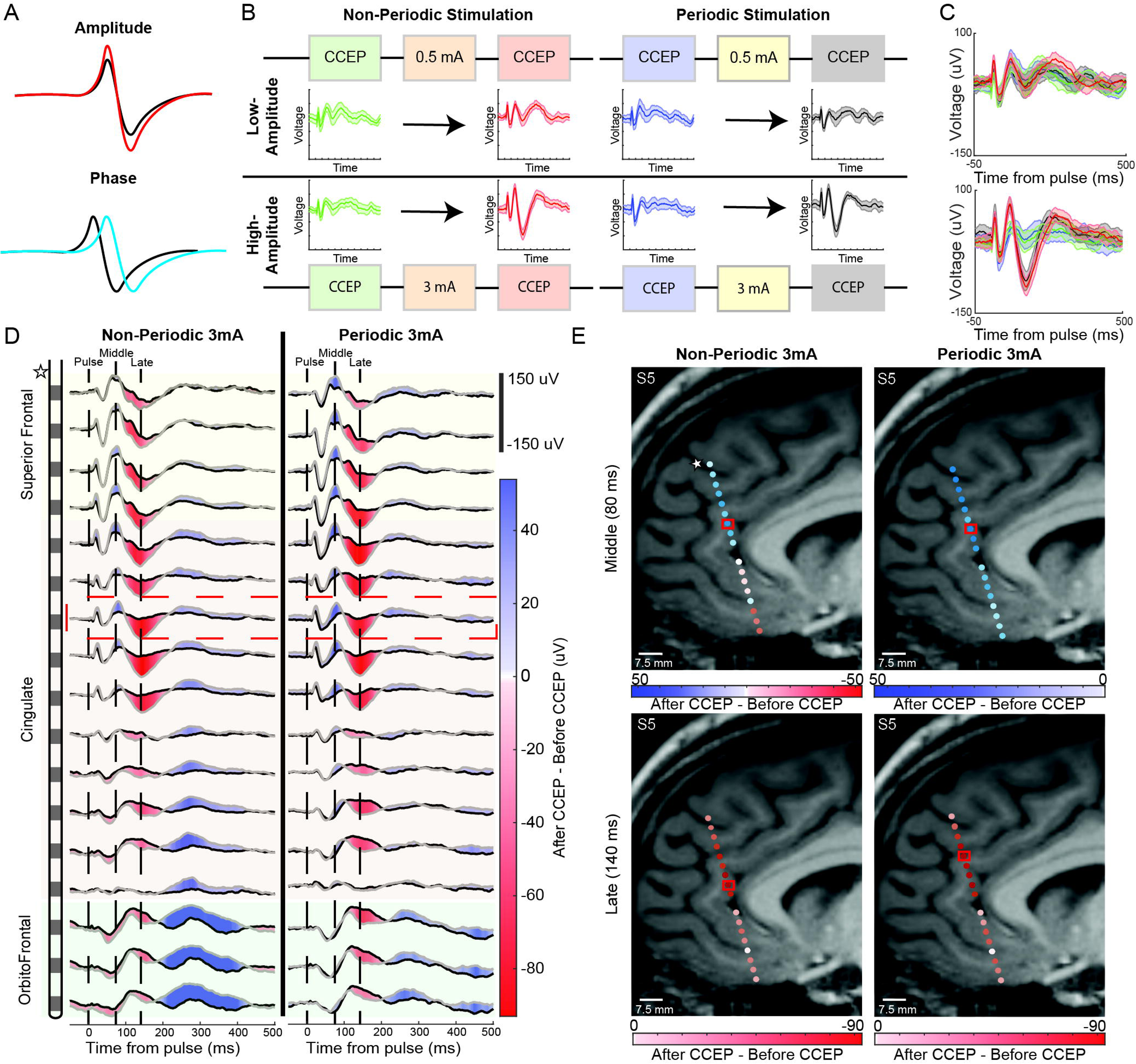
Modifications to a thalamic single-pulse elicited CCEPs vary spatially and depend on stimulation parameters. **a**, Thalamic stimulation-mediated CCEP response modifications in amplitude (red) and latency (blue). **b**, Data from patient S5 illustrating the experimental design (colored blocks) and resulting CCEP traces at one representative cortical recording site (RAC). Low-amplitude (0.5 mA) and high-amplitude (3 mA) ANT stimulation was delivered using non-periodic (left) and periodic (right) stimulation patterns. Shaded areas indicate the 95% confidence interval; solid lines represent mean responses across trials. **c**, Overlaid responses show no significant differences at low current (top), but significant response divergence at high current (bottom). Trace colors correspond to conditions shown in panel **b**. **d**, Spatially mapped response modifications across an example SEEG trajectory traversing superior frontal, anterior cingulate, and orbitofrontal cortices for high-amplitude stimulation with non-periodic and periodic patterns. Each line represents responses recorded from one electrode contact along the trajectory. Background color indicates spatial location, and structure labels are on the left. Black and gray lines indicate mean responses recorded before (black) and after (gray) stimulation. Color gradients (red to blue) show the magnitude and direction of response differences (after minus before stimulation), with darker colors indicating larger modifications. Vertical dashed lines mark stimulus onset (0 ms) and two representative response peaks at approximately 80 ms (“middle positivity”) and 140 ms (“late negativity”). The red dashed box highlights the example channel shown in panels **b** and **c**. **e**, MRI-rendered orthogonal slices aligned with the SEEG electrode trajectory from patient S5. Slices show spatial distributions of response modifications for the middle positivity at 80 ms and late negativity at 140 ms relative to non-periodic and periodic stimulation at high amplitude. Colors indicate magnitude, consistent with panel **d;** red box indicates the largest absolute difference across all contacts.

**Fig. 3:**
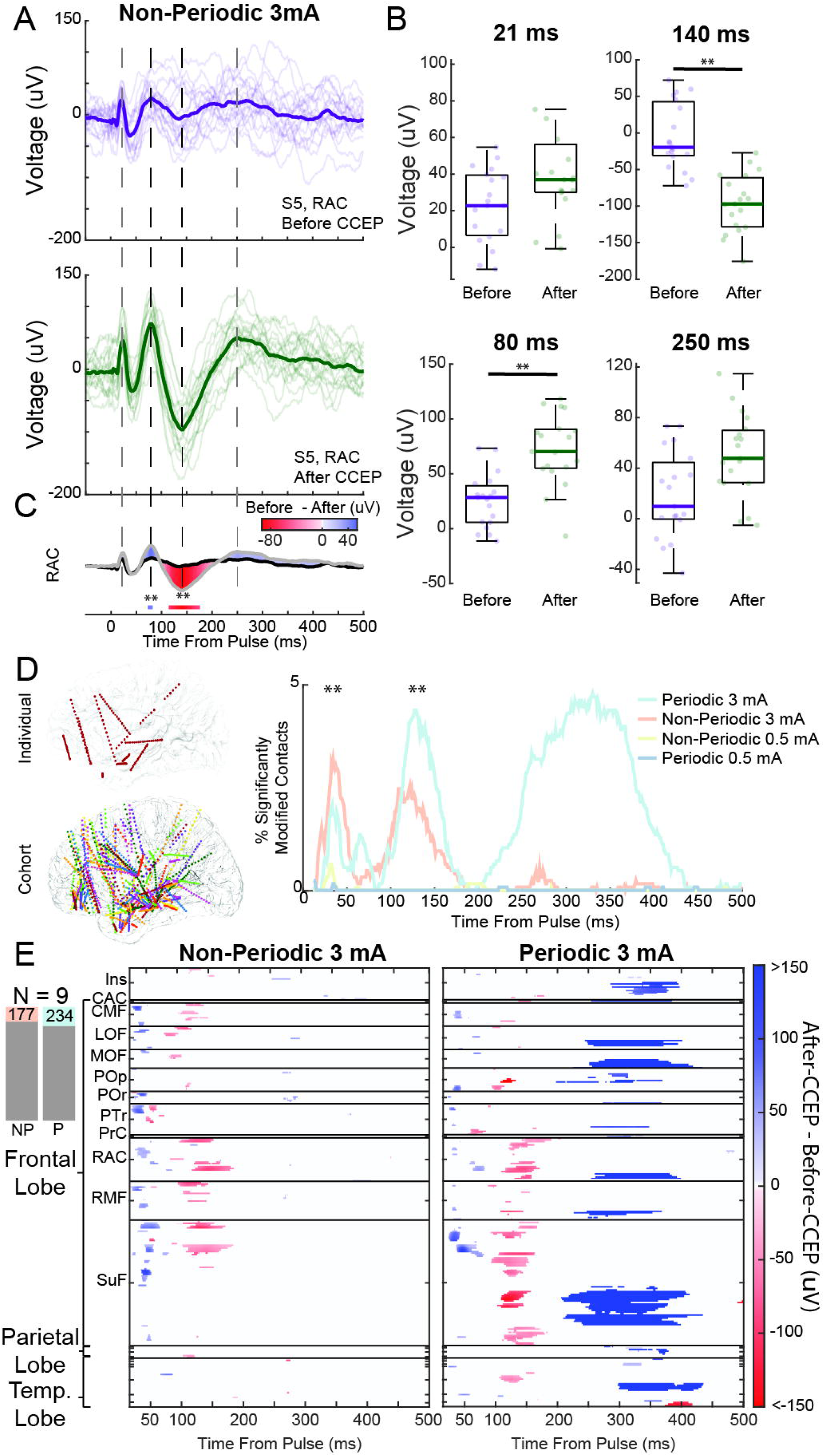
Significant CCEP modifications are spatially restricted to frontal structures and temporally limited to the first 150 ms post-pulse. **a**, Example single-trial CCEP responses recorded from the RAC in patient S5, before (purple) and after (green) non-periodic stimulation at 3 mA. Darker lines represent trial averages; lighter lines show individual trials. **b**, Box-and-whisker plots showing response amplitudes at four representative time points (21, 80, 140, and 250 ms). Central lines show medians; box edges indicate the 25th and 75th percentiles; whiskers extend to non-outlier values. Significant differences between conditions before vs after stimulation at 80 and 140 ms are indicated (\*\**P* < 0.0001, Bonferroni-corrected t-test). **c**, Overlay of tracing before and after stimulation tracings from panel **a** showing significantly different time points determined by t-tests exemplified in panel **b** (\*\**P* < 0.0001, Bonferroni-corrected t-test). **d**, Transition from individual patient data (patient S5) to cohort-aggregated data (patients S1–S9). Right panel shows the percentage of contacts with significant response modifications (*P* < 0.0001, Bonferroni-corrected t-test) across time (15–500 ms) for each stimulation condition, highlighting distinct time windows of maximal connectivity modification. (**P < 0.0001, t-test for significant differences between high-amplitude periodic versus non-periodic stimulation) **e**, Heatmap summarizing significant connectivity modifications across the patient cohort, grouped anatomically by FreeSurfer-defined brain structures and lobes. Only contacts with modified responses are shown. Bar plots (top left) indicate the number of contacts whose response was modified significantly in each stimulation condition. Ins, insula; CAC, caudal anterior cingulate; CMF, caudal middle frontal cortex; LOF, lateral orbitofrontal cortex; MOF, medial orbitofrontal cortex; NP, non-periodic; P, periodic; POp, pars opercularis; POr, pars orbitalis; PTr, pars triangularis; PrC, precentral gyrus; RAC, rostral anterior cingulate; RMF, rostral middle frontal cortex; SuF, superior frontal cortex.

To evaluate whether hemisphere or brain region influenced the likelihood of connectivity modifications we used generalized logistic mixed-effects models (GLMMs). With hemisphere as a fixed factor, the right-sided contacts had significantly higher odds of connectivity modifications than left-sided contacts (β = 0.556 ± 0.006, *P* < 0.00001). Separately, contralateral connectivity modifications were less likely than ipsilateral connectivity modifications (relative to the stimulated ANT) (β = −6.28 ± 0.05, *P* < 0.00001). These findings support that short-term ANT stimulation predominantly alters ipsilateral connectivity, consistent with strong intrahemispheric thalamic efferents^21^.

Next, we used lobe (frontal, parietal, temporal; occipital contacts removed due to low cohort sampling) as the fixed effect to model spatial specificity of response modifications. Irrespective of the stimulation pattern, the frontal lobe was most likely to show connectivity modifications (β = 1.876 ± 0.0062, *P* < 0.00001), whereas the parietal (β = −1.474 ± 0.0110, *P* < 0.00001) and temporal (β = −0.672 ± 0.0069, *P* < 0.00001) lobes had a lower propensity for modification.

We observed spatially restricted differences as an electrode transversed distinct structures, with components of the response being maximally different at distinct locations (Fig. 2d,e). We modeled the effects of structure (e.g., RAC, superior frontal, insula; refer to the DKTatlas for all segmented structures^47^) on the odds of observing connectivity modifications (see Fig. 3e for the occurrences of significantly modified responses per contact across all patients). Within the frontal lobe, the RAC (β = 3.303 ± 0.0086) and superior frontal cortex (β = 3.077 ± 0.0072) had the highest odds of showing connectivity modifications, followed by the caudal/rostral middle frontal cortex and lateral orbitofrontal cortex (all *P* < 0.00001). Temporal lobe subregions including the hippocampus and entorhinal cortex had lower odds of connectivity modification than other parts of the brain. Two non-frontal regions, insula and superior parietal cortex, had greater than average odds of connectivity modifications, though with smaller effect sizes than many frontal lobe structures (see full quantitative results in Supplementary Table 1).

Overall, these analyses indicate that high-current ANT stimulation robustly modifies thalamocortical connectivity, which are concentrated in the ipsilateral frontal lobe, especially the anterior cingulate and superior frontal cortex. Low-current stimulation elicited almost no measurable connectivity modifications. This spatial selectivity was consistent across patients and highlights that short-term ANT stimulation principally modifies the thalamo-frontal pathway.

### ANT stimulation induces temporally restricted response modifications

Although the clinical effects of ANT-DBS unfold over weeks to months, the immediate temporal profile of stimulation-induced network changes is undercharacterized. Thus, we compared the temporal profile of connectivity modifications due to the response. As previous studies have demonstrated CCEP response modifications occur 10–250 ms post-pulse^17,18^, we posited that short-term ANT stimulation would produce similar time-limited effects.

All segmented time bins had significant coefficients (*P* < 0.05), indicating a non-uniform effect of connectivity throughout the 15–500 ms response window (Fig. 3d, right panel). Effect sizes were generally largest for earlier bins (100–150 ms: β = 1.7751 ± 0.0041, *P* < 0.00001; 15–50 ms: β = 1.0117 ± 0.0051, *P* < 0.00001) and lower in later bins (350–400 ms: β = −0.2965, *P* < 0.00001; 450–500 ms: β = −4.0435, *P* < 0.00001; see full results in Supplementary Table 2). This pattern supports that most response modifications were concentrated in the first 150 ms. The 300–350 ms bin had a modest positive coefficient (β = 0.0582 ± 0.0044, *P* < 0.00001), suggesting a distinct increase in response modifications at that latency. This relatively small coefficient partly reflects a robust late response in one patient (S8) with a broad potentiation after high-amplitude periodic stimulation (Supplementary Fig. 3). In that patient, many contacts showed significant response modifications between 300–350 ms after periodic but not non-periodic stimulation. Because this effect is patient-specific, with response modification after 150 ms being rare for other patients, its overall contribution to the cohort-level model was limited. This analysis shows that short-term ANT stimulation primarily affects the first 150 ms after each pulse within our cohort.

### Short-term ANT stimulation drives selective response modifications in phase and amplitude in responsive brain regions

Within our cohort, we saw instances of response modifications altering latency without large amplitude-based response modifications (e.g., delayed or advanced peak) or modified amplitude while preserving overall timing (Fig. 2a). Thus, we hypothesized that multiple mechanisms may contribute to the connectivity modifications seen across the brain. To disentangle these contributions, we separated each high-current CCEP signal into instantaneous phase and amplitude components via the Hilbert transform. We first tested whether response modifications in phase or amplitude were randomly distributed across the 15–500-ms post-pulse window or clustered in specific intervals. Both phase (χ^2^ = 10553, df = 485, *P* < 0.00001) and amplitude (χ^2^ = 13303, df = 485, *P* < 0.00001) significantly deviated from uniformity. Thus, modifications in each phase and amplitude arose preferentially at certain latencies and were not uniformly scattered (see significant response modification peaks in distinct temporal segments Fig. 4f,g).

**Fig. 4:**
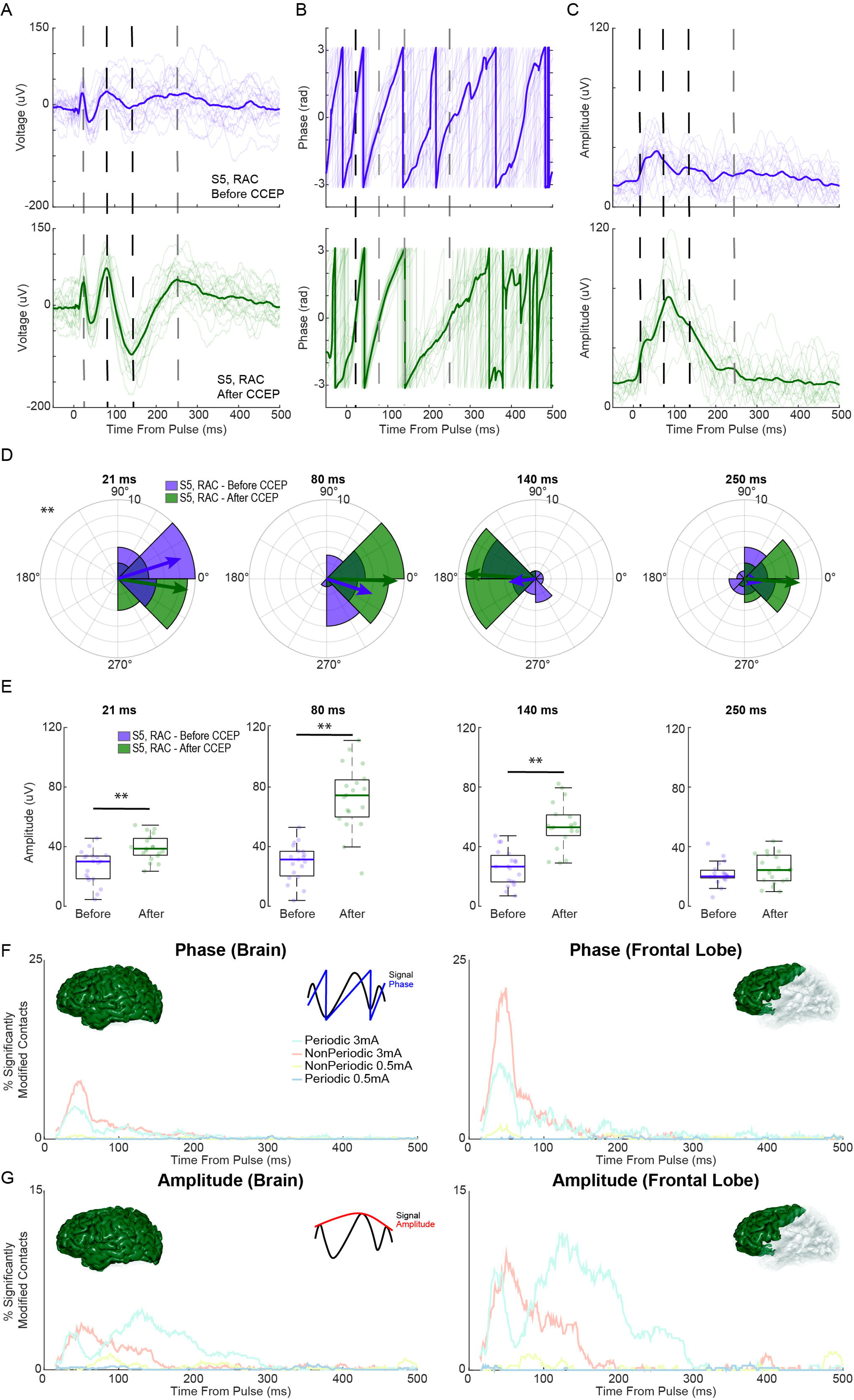
ANT stimulation preferentially modifies early CCEP response phases and later (< 51 ms) amplitudes. **a**, Example single-trial evoked response waveforms recorded from the RAC (patient S5) before (purple) and after stimulation (green). Dark lines represent mean responses; shading indicates individual trials. **b,c**, Phase (**b**) and amplitude (**c**) components corresponding to **a** were extracted using a Hilbert transform. Waveforms show stimulation-induced modifications to response timing and amplitude. **d**, Polar histograms illustrating phase distribution across trials at four representative time points (21, 80, 140, and 250 ms) from the data shown in **b**. Arrows indicate mean resultant vectors, with statistical significance determined by Kuiper tests (*P* < 0.001). **e**, Box plots comparing amplitude distributions at matched time points to **d** from the data shown in **c**, highlighting significant differences (\*\**P* < 0.0001, paired t-tests). Box plots show medians, interquartile ranges, and whiskers extending to non-outlier values. **f,g**, Cohort analysis showing the percentage of significantly modified responses for phase (**f**) and amplitude (**g**) over time (15–500 ms). Data are shown separately for brain-wide and frontal-lobe-only contacts after different stimulation conditions (periodic vs non-periodic; low vs high amplitude). Insets illustrate how phase and amplitude were quantified.

To compare the relative prevalence of phase vs amplitude response modifications at different latencies, we subdivided the response into three segments: early (15–50 ms), middle (51–150 ms), and late (151–300 ms). We then determined whether phase or amplitude response modifications were more frequent in each segment with McNemar tests. Phase-based response modifications dominated the early window (χ^2^ = 813.88, *P* < 0.00001), indicating that short-term ANT stimulation primarily shifted the timing of the earliest CCEP components. By contrast, amplitude-based response modifications were more common in the middle (χ^2^ = 1367.3, *P* < 0.00001) and late (χ^2^ = 821.24, *P* < 0.00001) windows (see the divergence of phase and amplitude curves at different latencies in Fig. 4f,g).

To further quantify the relative contributions of phase and amplitude, we modeled both characteristics as predictors of whether a channel’s evoked response was modified. Both phase and amplitude were significant predictors across the 15–500-ms window (*P* < 0.00001). This result is intuitive because both characteristics are simple decompositions of the original signal. However, this result shows that both amplitude and phase contribute to the response modification rather than being solely a phase or amplitude phenomenon.

To assess time-specific effects, we again subdivided the response window into the early (15–50 ms), middle (51–150 ms), and late (151–300 ms) intervals. We found that the Akaike Information Criterion (AIC) was lower for the phase-only model in the early interval (AIC_Phase_ = 9739, AIC_Amp_ = 11612). In the middle and late intervals, amplitude better explained the response modifications (middle: AIC_Phase_ = 38290, AIC_Amp_ = 35303; late: AIC_Phase_ = 40863, AIC_Amp_ = 34343). This analysis clarifies that short-term ANT stimulation initially alters the timing of the earliest cortical responses, with amplitude-based modifications emerging within the 51–150 ms response window.

To more easily interpret what is modified in the response in the two regions most affected by stimulation (superior frontal cortex and anterior cingulate), we estimated the timing change and gross potentiation with dynamic time warping (DTW) and an average amplitude metric (see Quantifying stimulation-induced amplitude potentiation and timing lag in the Methods). The regional specific timing markedly differed in the non-periodic vs periodic condition (Fig. 5a). In terms of the relative amplitude of the response, the majority of contacts within these two regions tended to be potentiated with both stimulation patterns (Fig. 5b).

**Fig. 5:**
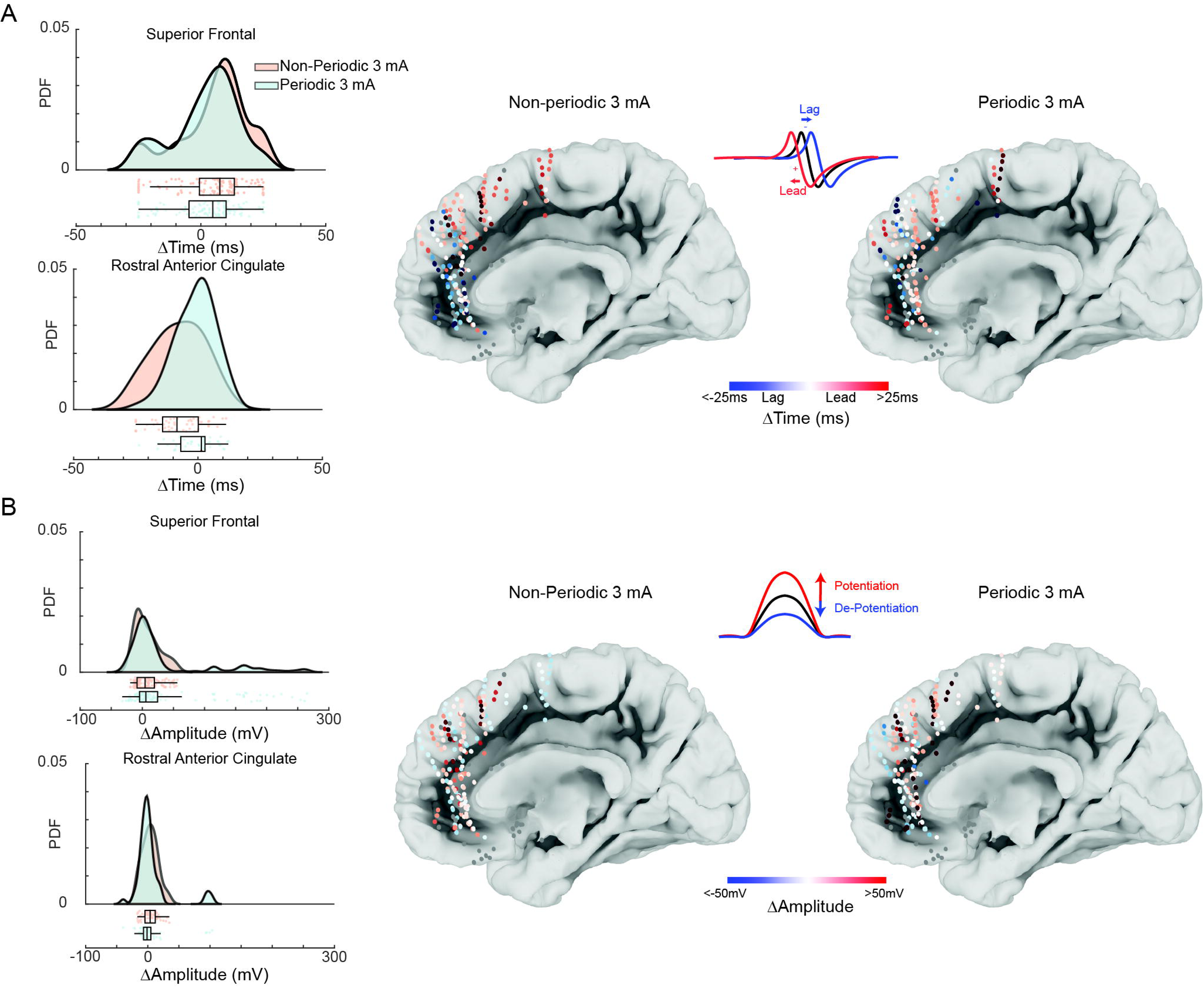
Potentiation and timing modifications of anterograde thalamic connectivity after short-term stimulation are spatially distinct. **a**, Channel-wise DTW-derived latency shift at 45 ms for every ipsilateral RAC and superior frontal contact. Negative values indicate that the response after stimulation lags the response before stimulation. The raincloud plot (left) summarizes the distribution of per-contact lags for non-periodic and periodic stimulation. PDF, probability density function. **b**, Channel-wise amplitude potentiation, defined as the average mean amplitude from 15–300 ms. Spatial map and raincloud plot are shown in the same format as **a**. Positive values denote net potentiation.

### Non-periodic stimulation of the ANT preferentially modifies early response components in connected frontal lobe subregions

In biophysical models, fluctuating frequency periods that are characteristic of non-periodic stimulation dynamically engaged networks and activated diverse neuronal populations^37^. Because non-periodic stimulation can engage with a greater range of different neuronal subtypes, we hypothesized that non-periodic stimulation would have a greater probability of affecting connected structures.

We examined whether the stimulation pattern engaged different brain regions (i.e., spatial specificity). We modeled this relationship with an interaction term between stimulation pattern and brain lobe (while controlling for ipsilateral classification), which significantly improved model fit (χ^2^ = 11364643, *P* < 0.00001). At the lobar level, non-periodic stimulation resulted in greater connectivity modifications in the frontal lobe (β = −0.51898, SE = 0.01389, *P* < 0.00001), whereas periodic stimulation resulted in greater connectivity modifications in the parietal (β = 0.09523, SE = 0.02540, *P* = 0.0002) and temporal (β = 2.0617, SE = 0.02167, *P* < 0.00001) lobes. We added an interaction term between stimulation pattern and individual structures to assess whether structures diverged with modified connectivity when accounting for pattern (χ^2^ = 12455835, *P* < 0.00001). Non-periodic stimulation had a greater likelihood of connectivity modifications than periodic stimulation in many frontal areas (e.g., caudal middle frontal cortex, rostral middle frontal cortex, superior frontal cortex, lateral orbitofrontal cortex, medial orbitofrontal cortex, pars triangularis, RAC), except for the pars opercularis.

Conversely, periodic stimulation had a greater likelihood of connectivity modifications than non-periodic stimulation in the temporal lobe (e.g., middle temporal cortex, superior temporal cortex, parahippocampal gyrus; see full results in Supplementary Table 3). To quantify how stimulation pattern influenced response timing, we modeled connectivity modifications across sequential time bins after short-term high-current stimulation. Non-periodic stimulation more strongly modulated connectivity in the earliest response window (15–50 ms: β = −0.73125, SE = 0.01354, *P* < 0.00001; Supplementary Table 4). However, in later windows, periodic stimulation predominantly drove response modifications, with progressively larger β coefficients. Although both patterns were effective, only the first 150 ms showed significant effects associated with lower standard errors, highlighting that the early segment is the most temporally discriminative between stimulation patterns (Fig. 3d).

To determine whether stimulation pattern resulted in different phase or amplitude response modifications, we modeled the interaction between stimulation pattern and phase/amplitude response modifications (χ^2^ = 1246.27, *P* < 0.00001). After non-periodic stimulation, phase modifications co-occurred more often with response modifications (phase: n = 6212, amplitude: n = 4771), whereas after periodic stimulation, amplitude modifications were more common (amplitude: n = 9220, phase: n = 4798). Consistent with this finding, non-periodic stimulation engaged phase shifts almost exclusively in the earliest response window (β_phase_ = 4.158 ± 0.082 vs β_amp_ = 2.382 ± 0.128; Wald F(1, 50,577) = 153.6, *P* < 0.008). In the middle window, both factors (β_phase_ = 2.672 ± 0.070; β_amp_ = 2.918 ± 0.069; *P* < 0.008) significantly predicted connectivity modifications, but neither factor dominated (Wald F(1, 140,497) = 6.2, *P* = 0.013). In the late window, only amplitude was significant (β_amp_ = 5.107 ± 0.140, *P* < 0.008; β_phase_ = –1.298 ± 1.024, *P* = 0.205; F(1, 210,747) = 37.7, *P* < 0.008). Under periodic stimulation, amplitude dominated both the early (β_amp_ = 4.433 ± 0.120 vs β_phase_ = 2.950 ± 0.131; F(1, 50,577) = 77.1, *P* < 0.008) and middle (β_amp_ = 3.627 ± 0.061 vs β_phase_ = 1.130 ± 0.088; F(1, 140,497) = 483.9, *P* < 0.008) period, and amplitude contributed equally in the late period (β_amp_ = 1.840 ± 0.045 vs β_phase_ = 2.006 ± 0.172; Wald F(1, 210,747) = 0.9, *P* = 0.35).

### Stimulation characteristics and recording contact location explain ANT-stimulated connectivity modifications

To characterize the factors associated with connectivity modifications mediated by short-term ANT stimulation, we compared the relative importance of features related to patients, anatomy, stimulation, and response. To identify significant predictors of connectivity modifications while reducing overfitting risk, we modeled using L1 regularization. This model excluded features that did not contribute significantly (*P* < 0.0007, Bonferroni-corrected for the number of initial regressors). The characteristics excluded were age, sex, distance from the stimulating dipole to the ANT-MTT junction, number of implanted contacts or trajectories, and stimulated hemisphere. Medication-related features (administration and timing relative to the experiment) also did not significantly covary with connectivity modifications. Connectivity modifications associated with the temporal lobe were generally nonsignificant, except with the middle (*P* < 0.00001) and superior temporal (*P* < 0.0041) cortices.

Next, we ranked retained features to evaluate their relative importance. The final model showed a strong overall fit (χ^2^ = 3.52 × 10^6^, df = 2,731,289, *P* <0.00001) with a McFadden Pseudo-R2 of 0.668. This finding indicates that the model accounts for more substantial deviance reduction than a null model. We further verified the contribution of individual predictors using delta deviance testing, which confirmed that each retained feature significantly improved model performance (Table 1, Fig. 6b). To understand how broader feature classes contributed to model performance, we grouped the surviving predictors into four categories by original classification:

1. Stimulation-specific factors: current amplitude, stimulation pattern, stimulation order, prior high-amplitude usage
2. Anatomy-specific factors: ipsilateral stimulation; frontal and parietal lobe labels; localization within the pars triangularis, pars opercularis, RAC, superior frontal cortex, superior temporal cortex, superior parietal cortex, middle temporal cortex, and insula
3. Patient-specific factors: patient ID (a unique identifier for each patient), time in the epilepsy monitoring unit (EMU), proximity of SOZ contacts to modified sites
4. Response-timing factors: time bins for response modifications spanning 15–500 ms post-pulse

**Fig. 6:**
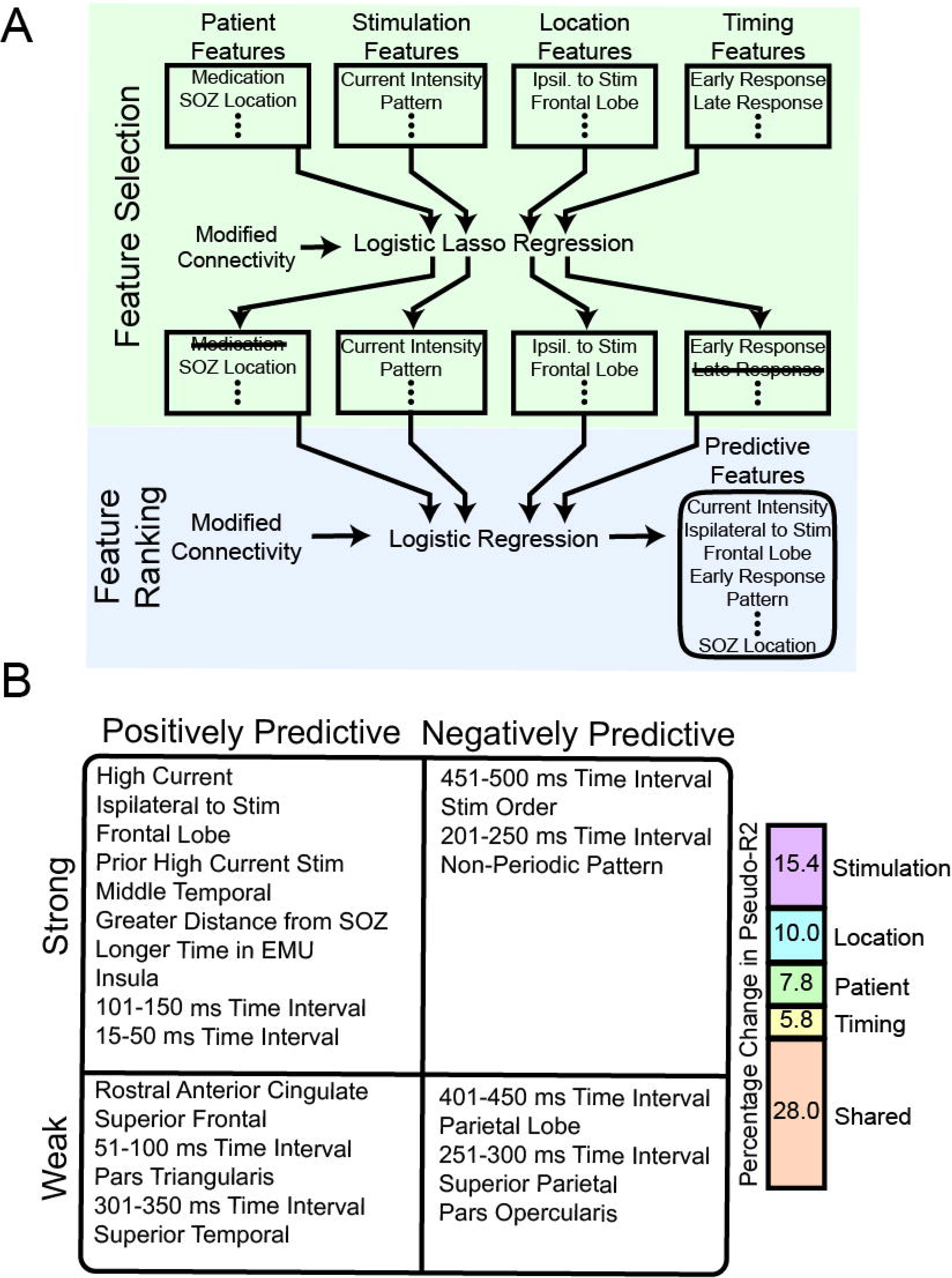
Stimulation and contact location factors dominate patient and timing of response factors when describing connectivity modifications. **a**, Schematic of the analysis pipeline: feature extraction, L1-regularized selection, and final logistic-regression ranking of predictors. **b**, Fixed-effect predictors ranked by odds ratio; strong vs weak predictive terms are further split into positively and negatively predictive associations. The bar plot (right) shows the proportional drop in Pseudo-R^2^ when each feature category is removed. “Shared’’ denotes variance jointly explained by more than one category.

**Table 1.**
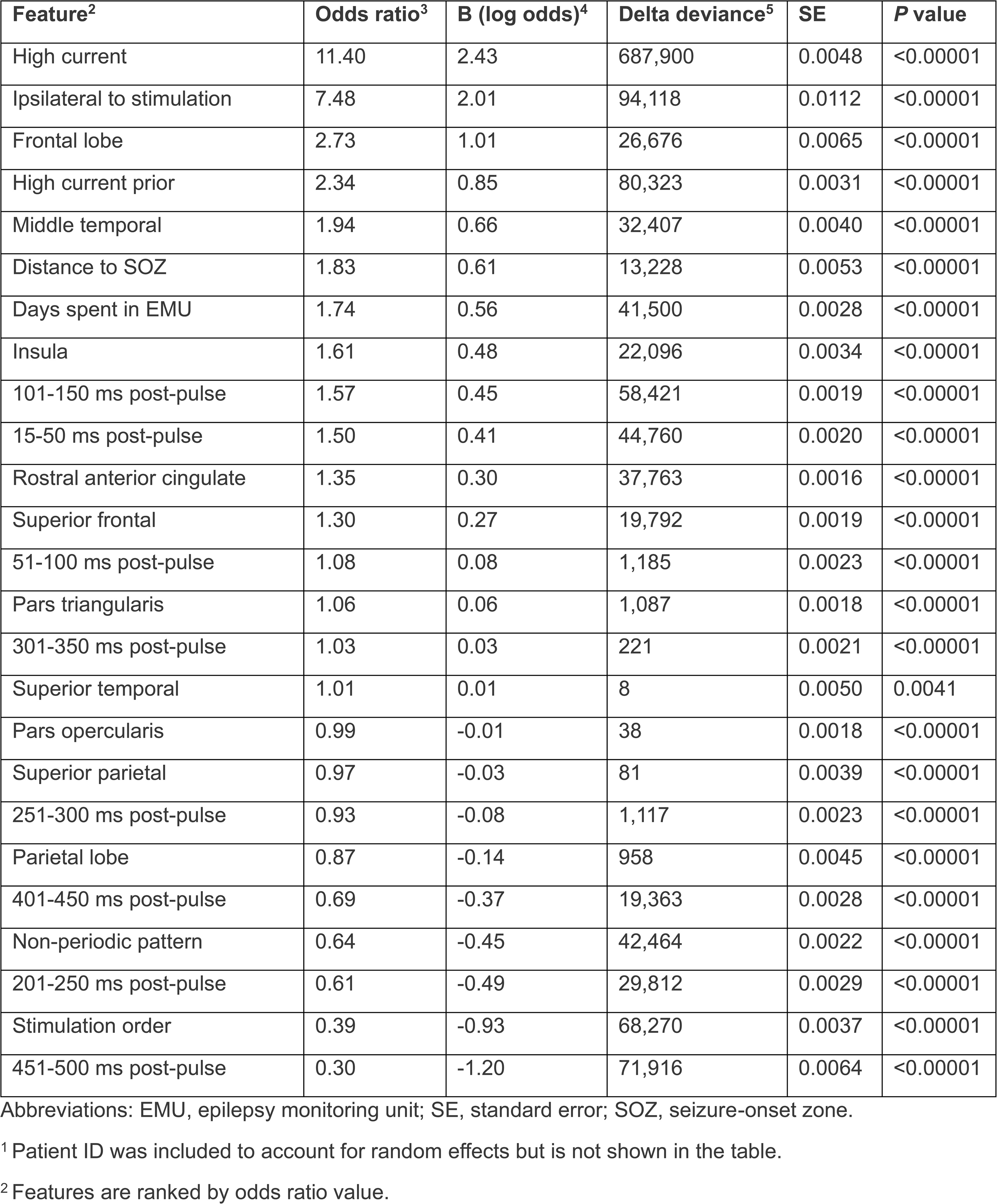
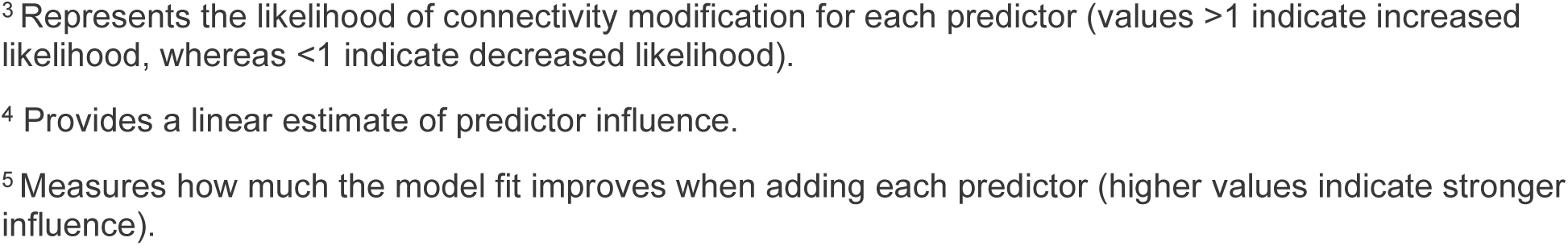
Generalized linear model results identify significant predictors of connectivity modifications after stimulation^1^.

| Feature<sup>2</sup> | Odds ratio<sup>3</sup> | B (log odds)<sup>4</sup> | Delta deviance<sup>5</sup> | SE | P value |
| --- | --- | --- | --- | --- | --- |
| High current | 11.40 | 2.43 | 687,900 | 0.0048 | <0.00001 |
| Ipsilateral to stimulation | 7.48 | 2.01 | 94,118 | 0.0112 | <0.00001 |
| Frontal lobe | 2.73 | 1.01 | 26,676 | 0.0065 | <0.00001 |
| High current prior | 2.34 | 0.85 | 80,323 | 0.0031 | <0.00001 |
| Middle temporal | 1.94 | 0.66 | 32,407 | 0.0040 | <0.00001 |
| Distance to SOZ | 1.83 | 0.61 | 13,228 | 0.0053 | <0.00001 |
| Days spent in EMU | 1.74 | 0.56 | 41,500 | 0.0028 | <0.00001 |
| Insula | 1.61 | 0.48 | 22,096 | 0.0034 | <0.00001 |
| 101-150 ms post-pulse | 1.57 | 0.45 | 58,421 | 0.0019 | <0.00001 |
| 15-50 ms post-pulse | 1.50 | 0.41 | 44,760 | 0.0020 | <0.00001 |
| Rostral anterior cingulate | 1.35 | 0.30 | 37,763 | 0.0016 | <0.00001 |
| Superior frontal | 1.30 | 0.27 | 19,792 | 0.0019 | <0.00001 |
| 51-100 ms post-pulse | 1.08 | 0.08 | 1,185 | 0.0023 | <0.00001 |
| Pars triangularis | 1.06 | 0.06 | 1,087 | 0.0018 | <0.00001 |
| 301-350 ms post-pulse | 1.03 | 0.03 | 221 | 0.0021 | <0.00001 |
| Superior temporal | 1.01 | 0.01 | 8 | 0.0050 | 0.0041 |
| Pars opercularis | 0.99 | -0.01 | 38 | 0.0018 | <0.00001 |
| Superior parietal | 0.97 | -0.03 | 81 | 0.0039 | <0.00001 |
| 251-300 ms post-pulse | 0.93 | -0.08 | 1,117 | 0.0023 | <0.00001 |
| Parietal lobe | 0.87 | -0.14 | 958 | 0.0045 | <0.00001 |
| 401-450 ms post-pulse | 0.69 | -0.37 | 19,363 | 0.0028 | <0.00001 |
| Non-periodic pattern | 0.64 | -0.45 | 42,464 | 0.0022 | <0.00001 |
| 201-250 ms post-pulse | 0.61 | -0.49 | 29,812 | 0.0029 | <0.00001 |
| Stimulation order | 0.39 | -0.93 | 68,270 | 0.0037 | <0.00001 |
| 451-500 ms post-pulse | 0.30 | -1.20 | 71,916 | 0.0064 | <0.00001 |

Abbreviations: EMU, epilepsy monitoring unit; SE, standard error; SOZ, seizure-onset zone.
<sup>1</sup> Patient ID was included to account for random effects but is not shown in the table.
<sup>2</sup> Features are ranked by odds ratio value.
<sup>3</sup> Represents the likelihood of connectivity modification for each predictor (values >1 indicate increased likelihood, whereas <1 indicate decreased likelihood).
<sup>4</sup> Provides a linear estimate of predictor influence.
<sup>5</sup> Measures how much the model fit improves when adding each predictor (higher values indicate stronger influence).

Removing stimulation-specific factors caused the largest drop in model fit (15.4% reduction in Pseudo-R2; likelihood ratio test [LRT] = 813,975, *P* < 0.00001), followed by anatomy-specific (10.0%; LRT = 525,686), patient-specific (7.8%; LRT = 413,517), and response-timing (5.8%; LRT = 306,446) factors.

We computed the residual portion of Pseudo-R2 not explained by any single group alone. We found that 28% of the model’s explanatory power likely reflects shared or interdependent effects between feature groups. Although Pseudo-R2 values are not strictly additive, this residual approximates the importance of interactions or overlapping contributions across predictor categories. These findings support that stimulation parameters combined with anatomic context predominantly drive connectivity modifications. Although patient-specific factors retain some contribution, features related to the stimulation protocol and spatial localization were the strongest determinants of altered connectivity.

## DISCUSSION

In this study, we evaluated a short-term biomarker of therapeutic response to short-term ANT-DBS stimulation in patients with drug-resistant epilepsy. We assessed CCEPs across the brain before and after 18 minutes of therapeutic ANT stimulation, which induced spatial and temporal modifications in evoked responses. Although the ANT has extensive anatomic connectivity across its three subnuclei^45,46^, the connectivity modifications were not observed to be uniformly distributed. Instead, they were primarily restricted to the frontal lobe, mostly within the superior frontal cortex and anterior cingulate. Most response modifications occurred within the first 150 ms post-pulse, with a local maxima within the first 50 ms. This specific latency is tied to more direct, single synaptic relay connectivity between two brain structures^48–50^. Low-frequency stimulation of the thalamus can enhance or recruit rhythmic brain activity, leading to increased synchronization of cortical signals, especially over the frontal lobe^51–54^. In our study, the stimulation frequency used to elicit CCEPs was lower (average, 0.2 Hz) than what typically produces the recruiting rhythm and is generally more associated with desynchronization^55,56^. Thus, although the modified connectivity we observed occurs in similar brain areas and over similar timescales, this connectivity is unlikely to be a direct consequence of that response.

The mechanism underlying seizure reduction mediated by ANT-DBS may involve neurotransmitter amounts^57–59^, apoptosis/neurogenesis of neuronal populations^60–62^, and protein expression^63,64^. One long-held theory is that neuromodulation mitigates seizure spread by disrupting network propagation^4^. Specifically, ANT-DBS works therapeutically by functionally “lesioning” ANT from the broader epileptic network^65,66^. By modulating ANT-frontal connectivity, stimulation may prevent seizure propagation from the mesial temporal lobe, reducing the likelihood of secondary generalization and clinical seizure expression^67^.

One prediction arising from the “lesioning” hypothesis is that stimulation suppresses activation of thalamo-frontal pathways to “isolate” the ANT. Contrarily, our results support enhanced thalamo-frontal connectivity after short-term ANT-DBS. One possibility for this difference is that there is a network-level mechanism by which ANT-DBS functions outside of lesioning ANT. Alternatively, ANT-DBS may stabilize thalamo-frontal connectivity related to the epileptic network, promoting a brain state less likely to propagate epileptic activity. Previous studies showed that epilepsy is associated with imbalanced connectivity, in which hyperactive epileptic networks disrupt normal, brain-wide communication^68–70^. In contrast, people without epilepsy show more distributed and functionally integrated connectivity patterns^71,72^. Also, repeated seizure activity may reinforce pathological network patterns, further increasing susceptibility to seizures^73–75^. By strengthening non-epileptic networks (i.e., those not commonly activated at the start or early propagation of seizures), ANT-DBS may help to restore a more balanced network state, decreasing susceptibility to generalizing seizures.

Our findings suggest that stimulation induces network plasticity, consistent with studies using noninvasive metrics of magnetoencephalography and fMRI for other disorders^76,77^. Such changes are consistent with long-term clinical benefits of DBS in epilepsy that can accumulate over years, and seizure rates do not immediately return to baseline after stimulation stops^2,78–80^. Plasticity can be categorized as short or long term, each with its own underlying mechanisms. We evaluated whether connectivity modifications reflect short- or long-term plasticity by examining recovery patterns between stimulation blocks. Our baseline period was 18 minutes, allowing for short-term recovery. In other studies of short-term plasticity within the thalamus (albeit within a different nucleus), high-frequency stimulation induced synaptic plasticity for up to 15 minutes^81^. Thus, we attempted to separate short-term and long-term plasticity by evaluating whether connectivity modifications were preserved between blocks (see example in Supplementary Fig. 4). Across the entire experimental window, connectivity modifications rarely persisted into subsequent stimulation blocks, suggesting that these modifications are short term. To further assess whether connectivity modifications carried over between stimulation periods, we incorporated stimulation order and prior high-current stimulation into our mixed-effects model. We found that later stimulation blocks reduced the likelihood of connectivity modifications, whereas prior high-amplitude stimulation increased the likelihood of connectivity modifications. Thus, although connectivity modifications measured by CCEPs are short-lived, prior modifications may prime the network for future changes. This notion aligns with the long-term DBS effects seen in patients with epilepsy, in which therapeutic benefit increased over time^2,3^.

To identify the mechanisms underlying short-term plasticity, we separated the responses into phase and amplitude components. We found that response modifications were strongly associated with phase shifts in the early response window, peaking at approximately 45 ms post-pulse. In contrast, significant amplitude response modifications were more prominent in the later response window (51–150 ms), often showing a prolonged or bimodal distribution. These findings suggest that response modifications in the earlier response components are predominantly related to timing rather than the strength of the response.

Our observed short-term plasticity in thalamocortical networks could be achieved by neurons and neural circuits through several mechanisms, including kinase-mediated connectivity modifications of channel kinetics or trafficking of additional ion channels to the synapse^82^. With DTW, we found a spatial relationship to response timing, with lagged responses occurring more often within RAC in the non-periodic condition. These results, coupled with a greater amplitude that co-occurred with these temporal shifts, support that ANT stimulation may modify frontal cortical channel kinetics. In previous studies with pain models, glutamate mediated the ability of intralaminar thalamic nuclei to induce short-term plasticity in anterior cingulate after electrical stimulation^83,84^. The long-term plastic effects could also be explained by kinase activation, which can enhance channel expression and receptor trafficking. This mechanism has been proposed to underlie transcranial magnetic stimulation^85^. In computational models of synaptic strength and heterogeneous excitatory networks, researchers described a greater likelihood of seizures when epileptic and non-epileptic networks become unbalanced^86^. Also, ANT-DBS modifies plasticity and gene expression in the mesial temporal lobe^64^. An alternative explanation is that the modified characteristics of the neuronal populations involved are occurring within ANT. Although connectivity modifications may partly occur in the ANT, the frontal lobe structures are likely being modified given the relative spatial specificity of the phenomenon. Despite this observation, we could not directly dissect the underlying mechanism in patients and therefore future studies in model organisms are needed.

A key finding of our study is that different stimulation parameters and patterns yield distinct connectivity modifications. For example, low-current stimulation rarely induced connectivity modifications, whereas high-current stimulation induced reliable changes both spatially and temporally. Because we use only two currents, we do not know the precise relationship between current strength and connectivity modifications. However, our results suggest that a minimum stimulation threshold is needed to drive circuit changes and that connectivity does not seem to fluctuate spontaneously over time. This suggestion is consistent with previous work^87^.

Although non-periodic stimulation produced fewer overall connectivity modifications than periodic stimulation, non-periodic stimulation induced stronger early phase-based response modifications and appeared to more effectively target direct thalamo-frontal pathways. In contrast, periodic stimulation produced broader, temporally extended potentiation. One possible explanation for this difference is that non-periodic stimulation encompasses a broader frequency spectrum that could prevent neuronal adaptation, increasing the likelihood of hitting an “optimal frequency” within the composite signal, and preventing hypersynchronization. This possibility aligns with rodent models of epilepsy, albeit at low stimulation frequencies^33^. Further, different stimulation frequencies can engage different proportions of neural populations^88^. In the ventral intermediate nucleus of the thalamus, adaptation to electrical stimulation can lead to worse clinical outcomes in patients with essential tremor^89,90^, which was overcome with varying stimulation parameters^91^. Because periodic stimulation effectively delivers tonic stimulation (around 145 Hz) with no variability, periodic stimulation may be more susceptible to network habituation or entrainment effects.

Our work shows clear differences in response profiles given the parameter set used to stimulate ANT. This readout is an attractive starting point to understand the utility of CCEPs in confirming target engagement and/or therapeutic effect. Our study does not directly correlate this biomarker with the clinical gold standard of reduced seizure rates, so we can only speculate what characteristics of the modified response might correlate with these rates. One possibility for how response modifications could predict seizure rates is that the higher gross number of instances of response modifications (as in periodic stimulation) is less important than affecting the earliest components of the response (as in non-periodic stimulation). Another possibility is the link between the success of stimulation in reducing seizures and the strengthening of connectivity outside the epileptic network^11^. The plastic effects we saw aligns conceptually with how seizure rates might be reduced long term by restructuring synaptic weights across the brain. This alignment supports examining these measures as potential guides for parameter tuning. This possibility has much broader implications than solely applications of ANT-DBS, because other neurological disorders (e.g., dystonia^92^, depression^93^), like epilepsy, require longer evaluations to gauge therapeutic effectiveness. This study was limited to studying connectivity modifications while the patient was in the EMU, thus it was not possible to define the relationship of connectivity modifications with long-term seizure rates. A follow-up study prospectively tracking connectivity modifications along with seizure rates utilizing minimally invasive electrodes across neuromodulation programming is currently being planned to draw this direct comparison.

When we evaluated other factors that covaried with connectivity modifications, we recognized that stimulation parameters and spatial specificity were more predictive of connectivity modifications than patient-related features (e.g., medication status, number of implants, seizure-onset location). However, patient variability is non-trivial. We precisely controlled for patient-specific effects in our mixed-effects models because of the degree of patient variability. For example, one patient (S8) responded strongly to high-amplitude periodic stimulation with a broad and late response modification, whereas another patient (S7) had relatively few response modifications. Thus, although stimulation parameters and anatomical context are dominant factors, patient-specific variation persists. To uncover how genetic, metabolic, or anatomic differences shape individual responses to stimulation, future studies with larger cohorts, more detailed longitudinal data, and large-animal models that recapitulate the human disease are needed.

In this study, we evaluated a candidate short-term biomarker of neuromodulation efficacy and provided crucial insights into the spatial and temporal dynamics of network plasticity driven by ANT stimulation. These insights advance our understanding of how subcortical stimulation modulates cortical connectivity, with immediate relevance to managing epilepsy. The mechanistic insights herein have broader implications for optimizing neuromodulation across neurologic and psychiatric disorders, ultimately guiding more effective therapeutic interventions and substantially improving patient care.

## ONLINE METHODS

### Patient demographics, selection, and ethical considerations

Nine adult patients (three male, six female; aged 19–65 years) with drug-resistant focal epilepsy who underwent intracranial EEG monitoring at Mayo Clinic Arizona were enrolled in the study and completed the experimental stimulation protocol. All patients provided written informed consent under a protocol approved by the Mayo Clinic Institutional Review Board (IRB #15-006530). The primary clinical indication for intracranial EEG monitoring was localization of seizure foci to evaluate candidacy for potential surgical intervention. Demographic and clinical details of patients are summarized in Supplementary Tables 5 and 6.

All enrolled patients received subcortical depth electrode implants that target the ANT as part of their standard Phase II surgical evaluation. Only patients who completed the entire stimulation and recording protocol without experiencing seizures during the study were included in the analyses.

As part of standard clinical procedures, antiepileptic drugs were tapered to facilitate seizure capture during intracranial EEG monitoring. Antiepileptic drug regimens were restored to stable therapeutic doses at least 11 hours (mean: 23 ± 14 hours) before starting the stimulation protocol. Patient admission to the EMU ranged from 5 to 18 days.

### Patient safety and monitoring

Throughout each patient’s involvement in the research stimulation protocol, intracranial EEG recordings were continuously monitored by a board-certified epileptologist, trained EEG technicians, and EMU nursing staff to promptly detect potential adverse effects or stimulation-induced seizures. Research technicians were present at all times to ensure any clinical changes or patient-reported discomfort were immediately identified and managed.

Before starting stimulation, the charge density delivered to patient tissue was calculated and confirmed to be below previously established safety thresholds^94^. Immediately after stimulation began, patient tolerance and comfort were closely evaluated, and any reported discomfort was documented and promptly addressed.

In the rare event that a seizure occurred during experimental stimulation, the electrical stimulation was immediately stopped upon confirmation of ictal activity. Then the experimental session for that patient was discontinued, and data collected from that session were excluded from all analyses.

### Electrode placement, localization, and labeling

The precise localization of SEEG electrode contacts was determined for each patient. Postoperative computed tomography (CT) images were co-registered with preoperative T1-weighted MRI scans using “Your Advanced Electrode Localizer” (YAEL) software^95^. This procedure used rigid-body transformations optimized by mutual information^95^. The alignment of imaging modalities was visually inspected in multiplanar views and refined manually when needed. This multi-step approach ensured accurate anatomical alignment, a crucial requirement validated by prior evaluations of SEEG localization methods. High-density artifacts on postoperative CT scans, corresponding to SEEG electrode contacts, were identified automatically within the YAEL software. Each contact’s centroid was then projected onto the co-registered MRI to enable precise anatomic labeling. The trajectories of implanted electrodes were reconstructed and overlaid onto each patient’s brain anatomy for verification by referencing anatomic landmarks and electrode coordinates to validate accuracy.

Each electrode was anatomically labeled using an automated segmentation process utilizing DKTatlas within FreeSurfer^47^. Previous work has described similar levels of accuracy in automated cortical labeling when compared to manual labeling^96^. We visualize the proportion of contacts sampled within each labeled structure (Fig. 1d).

### SOZ localization

Initial seizure localization was conducted by expert epileptologists during each patient’s admission to the EMU. Due to clinical rotations, the attending epileptologist varied across patients. However, all seizure localization findings were reviewed and confirmed at a weekly multidisciplinary epilepsy conference at least once per patient.

Before inclusion in the analysis, seizure-onset localization was independently re-assessed in a semi-blinded manner by at least one epileptologist. Epileptologists were provided three representative seizure recordings per patient, from which any identifying information or clinical notes had been removed to reduce potential bias. Complete blinding was not possible because patients were recently patients at Mayo Clinic Arizona whose clinical histories were known to the evaluating epileptologists. Nonetheless, this additional re-evaluation aimed to standardize and confirm SOZ determinations across the patient cohort.

### Electrophysiologic recordings

Patients were implanted with intracranial SEEG electrodes (DIXI Microdeep). Each patient received between 8 and 17 electrode trajectories, with the total contact count ranging from 141 to 250. Electrode placement was determined according to patient-specific seizure semiology and presurgical clinical evaluation criteria. Signals were amplified and recorded using either the g.HIamp 256-channel biosignal amplifier (g.tec) at a sampling rate of 4800 Hz, or the Arc Zenith 288-channel amplifier (Cadwell) at 2000 Hz. All recordings used a global reference (before re-referencing) with sub-galeal contacts contralateral to the suspected seizure focus as both the ground and reference. Although all responses focused on contacts recorded within gray matter structures, responses of white matter contacts were also recorded (Supplementary Fig. 2,3).

### Experimental protocol and stimulation

Each patient participated in an experimental protocol consisting of four pseudo-randomized stimulation blocks (Fig. 1c). Randomization was controlled to ensure roughly equal representation of each stimulation set within each order of occurrence. Each block comprised three segments: initial baseline, therapeutic stimulation, and post-therapy assessment. The baseline and post-therapy segments assessed modifications in functional connectivity, whereas the therapeutic segment delivered experimental stimulation.

All experimental stimulation targeted the junction between the ANT and MTT, which is hypothesized as an optimal site for seizure suppression^8^. Targeting accuracy was ensured by preoperative image fusion of patient-specific MRI and intraoperative CT scans using BrainLab neurosurgical planning software. Postoperative analyses included a multistep imaging workflow using FreeSurfer segmentation followed by manual refinement, with electrode locations further validated through YAEL software registration. The Euclidean distances between bipolar stimulation sites and the ANT-MTT junction were computed (Supplementary Fig. 1c). Post-implantation CT coordinates were transformed into the Montreal Neurological Institute space for consistency across patients.

Baseline and post-therapy segments consisted of 20 single-pulse biphasic stimulations (100 µs pulse width, 4 mA amplitude, 3–7 s interstimulus interval [ISI]). An 18-minute, no-stimulation baseline preceded each therapeutic stimulation segment to allow sufficient washout of short-term plasticity, as suggested by prior studies^81^.

The therapeutic segment involved continuous 18-minute stimulation without duty cycling at either high-current (3 mA) or low-current (0.5 mA) amplitudes. Two distinct stimulation patterns were tested: periodic stimulation (consistent 7 ms ISI [145 Hz]) and non-periodic stimulation (jittered 6–8 ms ISI [averaging 145 Hz, but varying between 125–165 Hz]).

### Data pre-processing Noise removal

All data manipulation and analyses were performed using MATLAB R2023b. Two primary sources of noise were removed before further analysis: stimulation artifacts and high-amplitude epileptiform activity. Stimulation artifacts (0–15 ms post-pulse) were replaced using validated artifact-removal methods that substituted artifact-contaminated segments with spectrally and temporally matched baseline data^97^. Trials contaminated by substantial interictal epileptiform activity were manually identified and excluded from subsequent analyses.

### Channel rejection, filtering, data segmentation, and re-referencing

All channels were individually inspected, and channels with electrical faults, artifacts, or anatomic misplacement were excluded. Also, channels located within 1 cm of stimulation dipoles were omitted to minimize artifact contamination.

A response modification was defined as a significant deviation in the response profile of a structure before vs after ANT stimulation. To analyze response modifications, data were bandpass-filtered between 0.5 and 300 Hz using a first-order Butterworth filter. To calculate instantaneous amplitude and phase without low-frequency bias, a narrower bandpass filter of 5–300 Hz was used. Line noise (60 Hz and harmonics) was eliminated using tenth-order narrowband notch filters (±1 Hz).

Data epochs were aligned to stimulation pulses and segmented from 500 ms pre-pulse to 2000 ms post-pulse. Median voltage from the pre-pulse baseline (500 ms) was subtracted to correct for residual DC offset.

Segmented data were re-referenced using Common Average Referencing with Local Adjustment^98^, which removed common variance based on the 15–300-ms post-pulse interval from 25% of channels, thus preserving evoked potential integrity (Fig. 1f).

### Modified CCEPs

CCEPs were computed as trial-averaged voltage traces within each experimental condition (Fig 3a). Response modifications were statistically tested at each 1-ms interval (15–500 ms post-pulse) using two-tailed t-tests comparing pre- and post-stimulation voltages (Fig 3b). Multiple comparisons were corrected with Bonferroni adjustments (*P* < 0.05/C, where C = 485). The mean CCEP waveforms and their 95% confidence intervals were calculated, and differences before vs after stimulation were visualized using colorimetric maps (Fig. 3c). Significant response differences were aggregated by anatomical structure using FreeSurfer-derived labels (Fig. 3e and Supplemental Fig. 2).

To capture non-linear dynamics in the evoked response, we segmented the post-pulse window (15–500 ms) into nine sequential 50 ms bins (15–50, 51–100, . . . , 451–500 ms). We measured the evoked potential of each contact (before vs after stimulation) within each bin, using only high-current conditions (3 mA). We then fitted a new GLMM incorporating these time bins as fixed factors. This approach distinguished whether connectivity modifications were specifically associated with particular latency ranges, while controlling for interpatient differences.

To visualize at the network level, we calculated the proportion of contacts exhibiting significant instances of modified response within the response window to identify intervals of increased significance across the entire brain.

Finally, we assessed whether different stimulation parameters yielded varying degrees of modified connectivity using a chi-square test of independence. Specifically, we compared how often high-current vs low-current stimulation produces connectivity modifications based on the recorded CCEPs.

### CCEP instantaneous amplitude and phase response modifications

Filtered signals (Fig. 4a) were decomposed into phase components (Fig. 4b) and instantaneous amplitude (Fig. 4c) via Hilbert transform, facilitating separate assessments of each component’s contribution to response modifications.

Phase differences were visualized using compass plots and mean resultant length vectors (Fig. 4d). Statistical significance for phase response modifications was assessed with Kuiper’s test (*P* < 0.001). Amplitude differences were evaluated using identical statistical and visualization methods for the filtered signal (Fig. 4e).

We analyzed the proportion of significantly modified responses by cortical lobe (frontal, temporal, parietal) (Fig. 4f,g). To assess whether amplitude or phase modifications dominated in specific response windows (15–50 ms, 51–100 ms, 101–200 ms), chi-square and McNemar tests were performed.

A concern with phase-based metrics is that higher signal-to-noise ratios (SNRs) can yield more stable phase estimates, potentially confounding interpretations of connectivity modifications^99^. To mitigate this concern, we implemented several methodologic safeguards. First, we performed all analyses as within-contact, within-patient comparisons of evoked responses before and after stimulation, thereby eliminating interelectrode confounds. Second, we re-referenced signals using the Common Average Referencing with Localized Adaptive weighting, which reduces shared noise and improves spatial specificity. Third, we manually excluded epochs containing large motion artifacts or epileptiform transients to avoid contamination of phase estimates. Importantly, we observed that phase shifts were most prominent early in the response (15–50 ms) and amplitude response modifications occurred in a much broader window. This temporal dissociation supports that phase-based response modifications are not merely a byproduct of enhanced SNR.

To assess the relative contributions of amplitude and phase to response modifications, amplitude and phase response modifications were incorporated into a weighted generalized linear model. Model fits were evaluated with LRT and AIC. The AIC balances model fit against model complexity, while penalizing unneeded parameters. A lower AIC indicates that a predictor set more efficiently explains the data. To test whether phase or amplitude predominately drives response modifications, we performed Wald χ² contrasts on the difference between the phase and amplitude coefficients within each of three a priori temporal windows (early: 15–50 ms; middle: 51–150 ms; late: 151–300 ms). A Bonferroni correction (α = 0.008) was applied to control for multiple testing across the three windows.

### Mixed-effects modeling and interaction effects

Multiple weighted GLMMs were used to assess the importance of various features in explaining the variance of connectivity modifications. GLMMs are an extension of logistic regression that allows both fixed effects (predictors of interest, such as hemisphere or brain region) and random effects (accounting for interpatient variability). We used this approach because each patient contributed multiple electrode contacts, so not accounting for patient-level correlations could inflate Type I error. By including patient ID as a random intercept, the model can account for patient-specific baselines.

We applied weights to emphasize connectivity modification during model fitting, reflecting its occurrence at roughly a 1:200 ratio relative to non-modification. In each model, patient ID was a random effect to control for per-patient variability within the cohort, whereas the fixed effects varied depending on the specific analysis. For each model, we reported the log odds, standard errors, and *P* values for each fixed effect after model convergence.

To determine interaction effects, we created an additional weighted generalized linear model that included interaction terms between the relevant fixed effects. We then compared this interaction model to a simplified model without interaction terms using a chi-squared test to assess relative fit.

We tested fixed-effects models related to spatial specificity (contact hemisphere, contact lobe, and contact structure; ANT stimulation preferentially modifies thalamo-frontal connectivity) and temporal specificity (ANT stimulation induces temporally restricted connectivity modifications). Interaction models primarily analyzed pattern-specific effects on spatial specificity, temporal specificity, and the underlying phase and amplitude mechanisms (non-periodic stimulation of the ANT preferentially modifies early response components in connected frontal lobe subregions).

### Feature selection, ranking, and categorization

To isolate the key predictors involved in modifying functional connectivity, we evaluated a set of parameters related to stimulation, patient characteristics, electrode location, and response timing to assess their covariance with connectivity modifications. Specifically, we considered patient age; sex; handedness; anti-seizure medications commonly prescribed across multiple patients; time from last ictal onset; time in the EMU; time from last medication dose; the number of implanted trajectories; the number of implanted contacts; the hemisphere, lobe, and structure of the contact; the distance to another contact identified as a SOZ; the distance to another contact being stimulated; the hemisphere being stimulated; the stimulation pattern; current intensity; stimulation order; whether a previously used high-stimulation block was used earlier in the experiment; and the time from the stimulation pulse within the response curve.

All categorical variables (i.e., sex, commonly administered medications, hemisphere, lobe, contact structure, stimulation hemisphere, stimulation pattern, and current intensity) were one-hot encoded. All numerical variables were standardized using z-scores to enhance model stability. Once encoded, features were selected using a weighted, cross-validated, L1-regularized, generalized linear model. The feature selection step using L1 regularization ensures that only significant features are included in the final comparison step^100^. Overfitting by including non-significant features could destabilize the model and reduce generalizability of the results. The same weights used in the mixed-effects models were applied, giving preference to connectivity modifications. Cross-validation was repeated ten times, applying the “one standard error” rule to determine the critical lambda value, thereby leading to a reduced feature set. This set was further refined by including only features with significance values below a Bonferroni-corrected threshold based on the original number of features (*P* < 0.0007).

After feature selection, we constructed a weighted generalized linear model with the remaining features to assess their relative importance in explaining connectivity modification. Odds ratios, log odds, standard errors, and *P* values were calculated for each feature. Features with an odds ratio ≥ 1.5 or ≤ 0.67 were deemed “strong” predictors, whereas features with an odds ratio > 1 were deemed “positive” predictors. Finally, we removed each feature individually and refit the model to measure each feature’s delta deviance from the full model (Fig. 6a).

### Quantifying stimulation-induced amplitude potentiation and timing lag

For the analyses in Figures 5a and 5b, we restricted the dataset to contacts that were (1) ipsilateral to the stimulated ANT lead and (2) anatomically labeled as RAC or superior frontal cortex—the two structures that exhibited the highest incidence of connectivity modification.

For each channel, we computed the analytic signal of the single-pulse response by using the modulus to obtain the instantaneous amplitude envelope A(t) and averaging this envelope over 15–300 ms after the stimulus:

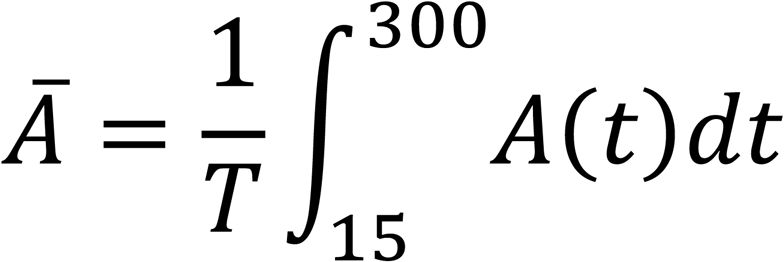

The relative potentiation was defined as 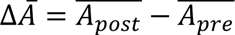, such that positive values indicate a net increase in response strength after the 18-minute stimulation block.

Latency shifts were quantified with a smoothed DTW approach applied to the raw CCEP traces averaged over trials. Responses after stimulation were aligned to their corresponding responses before stimulation within the 15–500 ms window using MATLAB’s DTW routine with a warping radius of 25 ms. For each channel, the resulting warping path provided a mapping *f*(*i*) from index *i* in the pre-trace to an estimated index in the post-trace. Duplicate indices in *f* were collapsed by averaging, after which an interpolant *f_est_* was generated. The instantaneous lag (converted to ms) was obtained as:

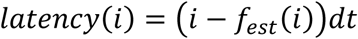

where *dt* is the sampling interval. Lags were smoothed with a 10-sample moving mean, and the value at 45 ms (the latency of maximal phase modulation identified in Fig. 4f) was extracted. Negative latency denotes a post-stimulation delay relative to pre-stimulation.

Channel-wise latency and potentiation values were displayed with a raincloud plot that overlays a kernel-density half-violin and box plot (Fig. 5a,b, left).

## Data Availability

All study data will be publicly available for use in Brain Imaging Data Structure format upon study peer-reviewed publication.

## Data and code availability

All study data will be publicly available for use in Brain Imaging Data Structure format upon study publication. All custom MATLAB functions used for processing data are available at: https://github.com/dbnl.

## Acknowledgments

This research was supported by funds from the Neurosurgery Research & Education Foundation. We offer gratitude to all our research subjects with drug-resistant epilepsy for their selfless participation in this study. We thank Crystal Herron, PhD, ELS(D), of Redwood Ink, LLC, for editing the manuscript.

## Author contributions

Teryn Johnson: Investigation, formal analysis, writing-original draft. Bobby Mohan: Investigation, data curation, visualization, writing – review & editing. Harvey Huang: methodology, writing – review & editing. Justin Cramer: Investigation. Cornelia Drees: Investigation, writing – review & editing. Matthew Hoerth: Investigation. Amy Crepeau: Investigation. Joseph Drazkowski: Investigation. Katherine Noe: Investigation. Leslie Baxter: Writing – review & editing. Amir Mbonde: Investigation. Christopher Harris: Investigation. Nuri Ince: Conceptualization. Kai Miller: Methodology, resources, software. Dora Hermes: Methodology, resources, software. Gregory Worrell: Conceptualization, resources, writing – review & editing. Jonathon Parker: Conceptualization, investigation, supervision, funding acquisition, writing – original draft.

## Competing Interests

Gregory Worrell is an inventor of intellectual properties developed at Mayo Clinic and licensed to Cadence Neuroscience Inc. and NeuroOne Inc. Mayo Clinic has received research support on behalf of Gregory Worrell from Cadence Neuroscience, UNEEG Medical, NeuroOne, and Medtronic. Gregory Worrell serves on scientific advisory boards for LivaNova, Neuropace, NeuroOne, and Cadence Neuroscience. No other authors believe they have competing interests as it pertains to the scientific discoveries within this manuscript.

## Notes

### Author Declarations

The IRB of Mayo Clinic gave ethical approval for this work (IRB #15-006530).

