## Supplementary figures and images for "Network Reorganization of Anterograde Thalamic Connectivity During Non-Periodic Patterned Deep Brain Stimulation"

### Supplemental Figure 1

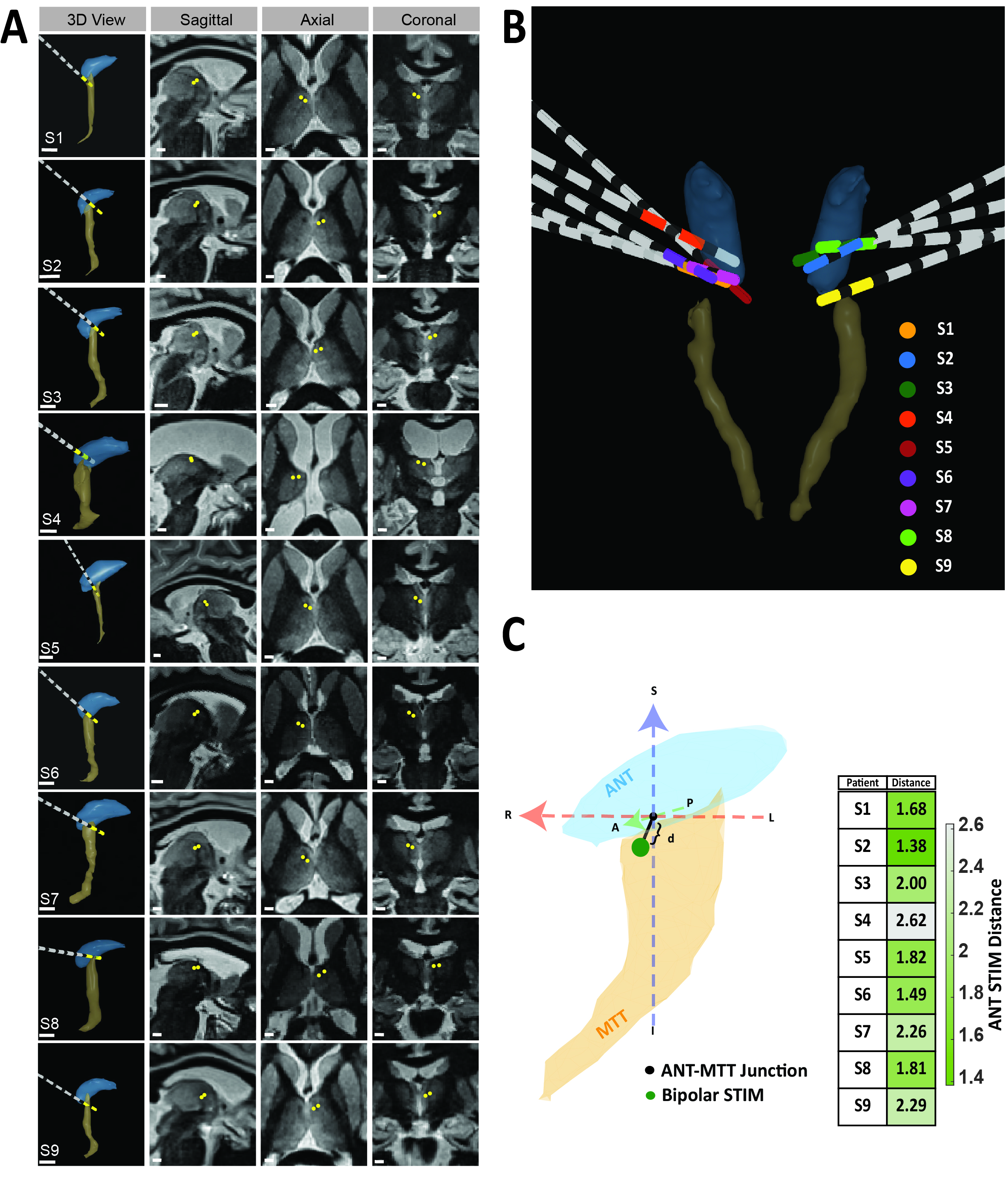

### Supplemental Figure 2

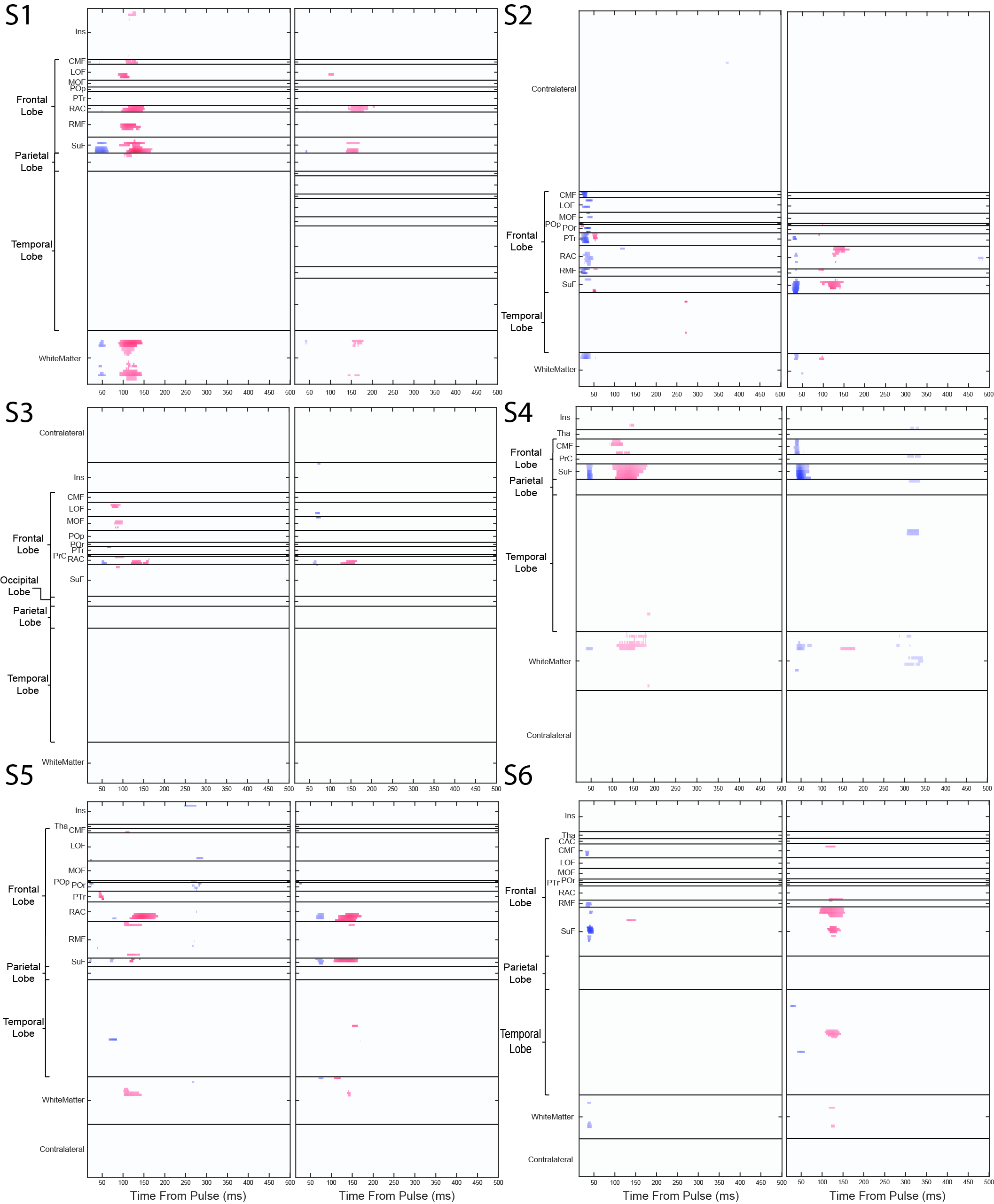

### Supplemental Figure 3

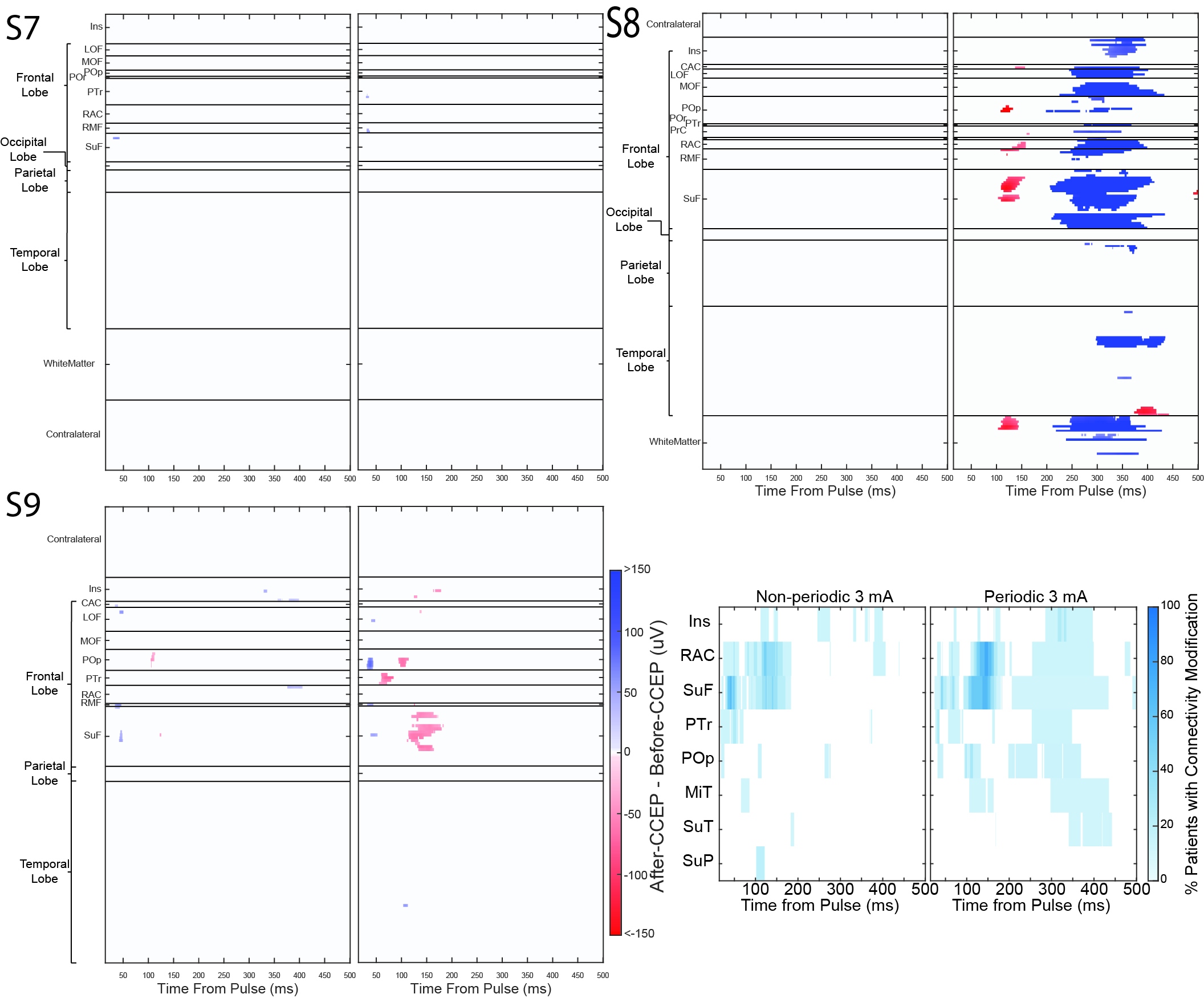

### Supplemental Figure 4

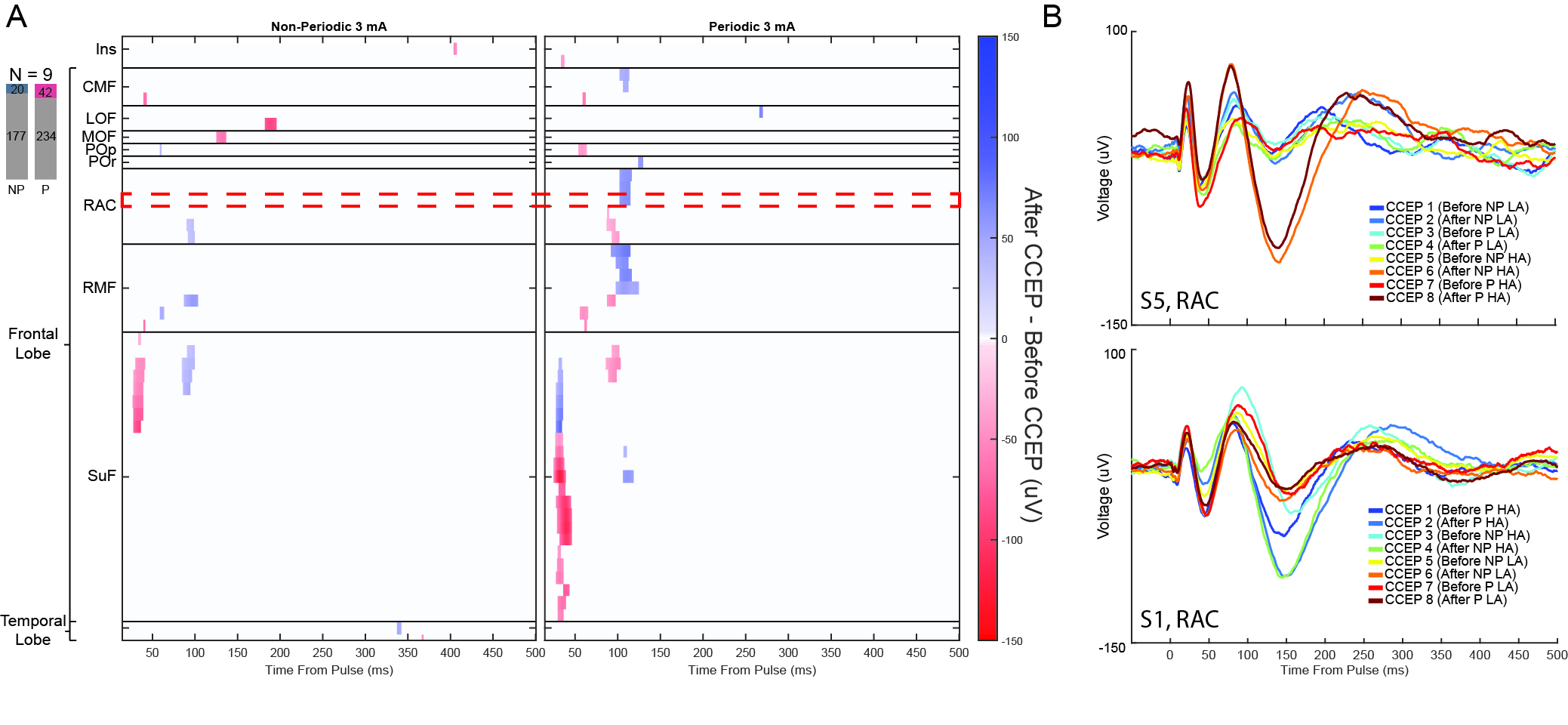
