## Supplemental Figure Captions for "Network Reorganization of Anterograde Thalamic Connectivity During Non-Periodic Patterned Deep Brain Stimulation"

**Supplementary Fig. 1: Target engagement of the SEEG electrode stimulating dipole with the ANT-MTT junction.** **a**, Patient-specific visualization of stimulation target engagement relative to the ANT-MTT junction. The ANT and MTT regions were segmented using the Thomas atlas from T1 MRI and further refined with expert guidance. Each two-dimensional slice (sagittal, coronal, and axial) depicts the bipolar stimulation contacts (yellow) overlaid on the fGATIR MRI image. **b**, Consolidated visualization of stimulation target engagement across all nine patients, rendered in the Montreal Neurological Institute (MNI) template space. Bipolar stimulation contacts were transformed from patient-specific coordinates to MNI template coordinates using Advanced Normalization Tools and are color-coded to distinguish patients. **C**, Distance metrics between the bipolar stimulation center and the ANT-MTT junction. The Euclidean distance was calculated for each patient (right).

**Supplementary Fig. 2: Significant responses across time and brain structures.** Each panel represents a different patient (S1–S6). Left, high-amplitude non-periodic stimulation; right, high-amplitude periodic stimulation. Gray lines indicate structural boundaries, with white matter contacts included. Shading shows significant increases (red) and decreases (blue).

**Supplementary Fig. 3: Significant responses across time and brain structures.** Each panel represents a different patient (S7–S9). Left, high-amplitude non-periodic stimulation; right, high-amplitude periodic stimulation. Gray lines indicate structural boundaries, with white matter contacts included. Shading shows significant increases (red) and decreases (blue). Bottom right: Heat map of the fraction of patients (not channel average) that show at least one significantly modified contact in each structure and time bin, displayed separately for high-amplitude non-periodic and periodic stimulation

**Supplementary Fig. 4: Persistence of potentiation beyond a single block.** **a**, Heatmap illustrating significant differences in pre-CCEP responses between consecutive blocks with high-amplitude non-periodic (NP) and periodic (P) stimulation. Color shading indicates regions of significant increase (red) or decrease (blue) in response amplitude relative to the before-CCEPs between itself and the next block. Ins, insula; CMF, caudal middle frontal cortex; LOF, lateral orbitofrontal cortex; MOF, medial orbitofrontal cortex; POp, pars opercularis; POr, pars orbitalis; RAC, rostral anterior cingulate; RMF, rostral middle frontal cortex; SuF, superior frontal cortex; **b**, Example CCEP waveforms showing different patterns of connectivity modification. Top, short-term connectivity change that does not persist into the next block; bottom, long-lasting connectivity change that continues into the next block. HA, high amplitude; LA, low amplitude; LTP, long-term potentiation.
