## Supplemental Tables for "Network Reorganization of Anterograde Thalamic Connectivity During Non-Periodic Patterned Deep Brain Stimulation"

**Supplementary Table 1.** Brain region-based GLMM modeling highlights frontal and cingulate predominance of ANT-stimulation-induced connectivity modification

| Cortical structure^1,2^ | Beta^3^ | SE | *P* value |
| --- | --- | --- | --- |
| Frontal lobe subregions |  |  |  |
| Rostral anterior cingulate | 3.3034 | 0.0086 | <0.00001 |
| Superior frontal | 3.0766 | 0.0072 | <0.00001 |
| Caudal middle frontal | 2.0933 | 0.0112 | <0.00001 |
| Rostral middle frontal | 2.0041 | 0.0093 | <0.00001 |
| Pars triangularis | 1.8832 | 0.0106 | <0.00001 |
| Pars opercularis | 1.4541 | 0.0106 | <0.00001 |
| Lateral orbitofrontal | 1.6202 | 0.0092 | <0.00001 |
| Medial orbitofrontal | 0.7436 | 0.0099 | <0.00001 |
| Temporal lobe subregions |  |  |  |
| Middle temporal | 0.9656 | 0.0082 | <0.00001 |
| Superior temporal | -0.9635 | 0.0010 | <0.00001 |
| Parahippocampus | -0.9568 | 0.0204 | <0.00001 |
| Fusiform | -0.7852 | 0.0294 | <0.00001 |
| Inferior temporal | -0.0390 | 0.0174 | <0.00001 |
| Hippocampus | -2.0663 | 0.0391 | <0.00001 |
| Other subregions |  |  |  |
| Insula | 0.0643 | 0.0092 | <0.00001 |
| Superior parietal | -1.7083 | 0.0082 | <0.00001 |

Abbreviations: ANT, anterior nucleus of the thalamus; GLMM, generalized logistic mixed-effects model; SE, standard error.

^1^ Each row indicates the estimated effect of that structure (relative to the model’s baseline category, typically “other structures” or “intercept”) on the likelihood of observing a significant cortico-cortical evoked potential modification.

^2^ Structures with too rare response modifications (e.g., amygdala entorhinal cortex, supramarginal cortex) were excluded due to non-convergence.

^3^ All values correspond to high-current (3 mA) stimulation only.

**Supplementary Table 2.** Connectivity modifications induced after ANT stimulation show time domain specificity

| Time bin, ms | Beta^1^ | SE | *P* value |
| --- | --- | --- | --- |
| 15-50 | 1.0117 | 0.0051 | <0.00001 |
| 51-100 | -0.0219 | 0.0052 | <0.00001 |
| 101-150 | 1.7751 | 0.0041 | <0.00001 |
| 151-200 | -0.0368 | 0.0050 | <0.00001 |
| 251-300 | -0.1779 | 0.0044 | <0.00001 |
| 301-350 | 0.0582 | 0.0044 | <0.00001 |
| 351-400 | -0.2965 | 0.0047 | <0.00001 |
| 401-450 | -1.6893 | 0.0068 | <0.00001 |
| 451-500 | -4.0435 | 0.0193 | <0.00001 |

Abbreviations: ANT, anterior nucleus of the thalamus; SE, standard error.

^1^ Positive values indicate a higher likelihood of modifications during that bin (relative to the model’s baseline); negative values indicate a lower likelihood.

**Supplementary Table 3.** Effects of stimulation pattern on individual brain structures

| Cortical structure | Beta^1^ | SE | *P* value |
| --- | --- | --- | --- |
| Frontal lobe subregions |  |  |  |
| Caudal middle frontal | -2.43000 | 0.02423 | <0.00001 |
| Rostral middle frontal | -2.09390 | 0.01946 | <0.00001 |
| Superior frontal | -0.34391 | 0.01629 | <0.00001 |
| Lateral orbitofrontal | -1.25610 | 0.02005 | <0.00001 |
| Medial orbitofrontal | -0.10257 | 0.02288 | <0.00001 |
| Pars triangularis | -1.61180 | 0.02194 | <0.00001 |
| Rostral anterior cingulate | -1.24890 | 0.01817 | <0.00001 |
| Pars opercularis | 0.070111 | 0.02720 | <0.00001 |
| Temporal lobe subregions |  |  |  |
| Middle temporal | 1.77090 | 0.02823 | <0.00001 |
| Superior temporal | 1.59010 | 0.03957 | <0.00001 |
| Parahippocampus | 1.00830 | 0.06400 | <0.00001 |
| Hippocampus | - | - | >0.05 |
| Inferior temporal | - | - | >0.05 |

Abbreviation: SE, standard error.

^1^ Positive values indicate a greater likelihood of connectivity change after periodic vs non-periodic stimulation; negative values indicate a greater likelihood under non-periodic vs periodic stimulation. All values were computed after high-current (3 mA) stimulation.

**Supplementary Table 4.** Non-periodic vs periodic stimulation selectively alters early anteriograde thalamic connectivity

| Time bin, ms | Beta^1^ | SE | *P* value |
| --- | --- | --- | --- |
| 15-50 | -0.73125 | 0.01354 | <0.00001 |
| 51-100 | 0.05500 | 0.01046 | <0.00001 |
| 101-150 | 0.64338 | 0.01066 | <0.00001 |
| 151-200 | 0.87173 | 0.01140 | <0.00001 |
| 201-250 | 4.74560 | 0.04472 | <0.00001 |
| 251-300 | 2.62610 | 0.02289 | <0.00001 |
| 301-350 | 6.29400 | 0.07592 | <0.00001 |
| 351-400 | 3.36100 | 0.02740 | <0.00001 |
| 401-450 | 3.60950 | 0.03055 | <0.00001 |
| 451-500 | 3.39140 | 0.02355 | <0.00001 |

Abbreviation: SE, standard error.

^1^ Positive values indicate a greater likelihood of connectivity change after periodic vs non-periodic stimulation; negative values indicate a greater likelihood after non-periodic vs periodic stimulation. All values were computed after high-current (3 mA) stimulation.

**Supplementary Table 5**. Patient demographics and clinical characteristics

| Patient ID | Age Range, y | Sex | Handedness | Onset Age Range, y | Seizure Type | PET^4^ | MEG | MRI | SISCOM | Prior Surgery |
| --- | --- | --- | --- | --- | --- | --- | --- | --- | --- | --- |
| 1 | 26-30 | F | R | 16-20 | Focal impaired awareness | L anteromedial, ant. temporal, bifrontal hypometabolism | NA | Normal | L temporal hyperperfusion | No |
| 2 | 36-40 | F | R | 11-15 | Focal bilateral to tonic-clonic | R prefrontal, sensorimotor, L occipital hypometabolism | NA | Normal | NA | No |
| 3 | 16-20 | F | R | 16-20 | Focal impaired awareness | R temporal hypometabolism | R Temporal | Normal | NA | No |
| 4 | 61-65 | F | R | 56-60 | Focal impaired awareness | L anterior medial temporal hypometabolism | NA | L mesial temporal ablation (prior sclerosis) | NA | Stereotactic ablation |
| 5 | 26-30 | M | R | 21-25 | Focal impaired awareness | Negative | NA | Normal | NA | No |
| 6 | 16-20 | M | A | 16-20 | Focal bilateral to tonic-clonic | Non-diagnostic | NA | R skull base encephalocele, L frontal venous anomaly | R temporal/frontal | No |
| 7 | 21-25 | M | R | 11-15 | Focal impaired awareness | L temporal hypometabolism | L Temporal | L anterior temporal ablation, encephalocele | NA | Yes |
| 8 | 51-55 | F | R | 31-35 | Focal Impaired Awareness | R Medial Prefrontal, Bilateral Posterior Cingulate, R Precuneus Hypometabolism | R Temporal/ Orbitofrontal | Small Bilateral Temporal Encephalocele | NA | No |
| 9 | 41-45 | F | R | 36-40 | Focal Bilateral to Tonic-Clonic | Negative | No Epileptiform Discharges | Normal | NA | VNS |

Abbreviations: A, ambidextrous; F, female; L, left; M, male; MEG, magnetoencephalography; MRI, magnetic resonance imaging; NA, not applicable; PET, positron emission tomography; R, right; SISCOM, subtraction ictal SPECT co-registered to MRI; VNS, vagus nerve stimulator.

**Supplementary Table 6.** Phase I and Phase II EEG findings, surgical interventions, and outcomes for each patient

| Patient ID | Phase I scalp EEG^1^ | Phase II intracranial EEG^2^ | Bilateral SEEG implant | Surgical intervention | Trajectories, No.^3^ | Contacts, No.^4^ | Stim hemi | LOS, days |
| --- | --- | --- | --- | --- | --- | --- | --- | --- |
| 1 | L temporal seizures | L mesial temporal seizures | No | L mesial temporal laser ablation | 12 | 179 | L | 5 |
| 2 | R temporal/post. temporal | Polyspikes in R/L mid/post. hippocampus | Yes | Bitemporal RNS | 13 | 211 | R | 18 |
| 3 | R temporal onset | R hippocampus head/mid | Yes | R mesial temporal laser ablation | 13 | 215 | R | 7 |
| 4 | L temporal seizures | L post. hippocampus, tail, residual amygdala | Yes | No subsequent intervention | 9 | 141 | L | 14 |
| 5 | L temporal seizures | L mesial temporal/neocortical structures | Yes | Bilateral ANT DBS | 12 | 196 | L | 11 |
| 6 | L frontal temporal | Broad L lateral temporal, orbitofrontal seizures | Yes | Bilateral ANT DBS | 8 | 236 | L | 16 |
| 7 | L temporal/post. onset | L mesial temporal structures | Yes | L ANT and L hippocampus RNS | 17 | 250 | L | 6 |
| 8 | R temporal | R hippocampus head, ant. sup. temporal gyrus | Yes | R temporal lobectomy | 17 | 233 | R | 9 |
| 9 | R temporal | R ant./mid. hippocampus, less from l hippocampus | Yes | R mesial temporal laser ablation | 17 | 195 | R | 11 |

Abbreviations: DBS, deep brain stimulation; ECoG, electrocorticography; EEG, electroencephalography; L, left; LOS, length of stay; R, right; RNS, responsive neurostimulation; SEEG, stereoelectroencephalography.

^1^ Non-invasive monitoring indicating seizure onset zones.

^2^ SEEG or ECoG specifying precise seizure origin within deep brain structures.

^3^ Number of implanted electrode trajectories during SEEG evaluation.

^4^ Total number of electrode contacts placed for SEEG monitoring.
